# A highly efficient T-cell immunoassay provides assessment of B cell help function of SARS-CoV-2 specific memory CD4^+^ T cells

**DOI:** 10.1101/2021.08.12.21261970

**Authors:** Asgar Ansari, Shilpa Sachan, Bimal Prasad Jit, Ashok Sharma, Poonam Coshic, Alessandro Sette, Daniela Weiskopf, Nimesh Gupta

## Abstract

The B cell help function of CD4^+^ T cells may serve as an immunologic correlate of protective adaptive immunity. The quantitative assessment of the B cell help potential of CD4^+^ T cells is limited by the lack of suitable antigen-specific functional assays. Here, we describe a highly efficient antigen-specific T-B co-cultures for quantitative measurement of T-dependent B cell responses. Using *Mycobacterium tuberculosis* specific setup, we show that early priming and activation of CD4^+^ T cells is important for the mutualistic collaboration between antigen-specific T and B cells, which could be achieved by supplementing the co-cultures with autologous monocytes. We further show that monocyte-derived growth factors provide the impetus for productive T-B collaboration by conferring optimal survivability in the cultured cells. This study provides first evidence of C-type lectin domain family 11 member A (CLEC11A/SCGF) as an essential growth factor for B cell survival. Importantly, we demonstrate the successful translation of monocyte supplemented T-B co-cultures in qualitative assessment of SARS-CoV-2 specific memory CD4^+^ T cells by quantifying several correlates of productive T-B cross-talk like plasma cell output, secreted antibody, antibody secreting cells and IL21 secreting T cells. Thus, the method described here can provides qualitative assessment of SARS-CoV-2 spike CD4^+^ T cells after natural infection and can be applied to assess the B cell help function of memory CD4^+^ T cells generated in response to COVID-19 vaccine.

**One sentence summary:** Qualitative assessment of antigen-specific CD4^+^ T cells for T-dependent B cell responses.

## Introduction

CD4^+^ T helper cells play crucial role in establishing the humoral arm of adaptive immunity. By supporting B cell function and antibody responses, CD4^+^ T cells play important role in controlling various infections caused by bacteria, fungus and virus. The CXCR5 expressing subset of CD4^+^ T cells known as follicular helper T (Tfh) cells are the specialized subset that provide help to B cells (Crotty, 2011). Tfh cells express CXCR5 to migrate to B cell follicles where they make cognate interactions with B cells and provide essential signals for survival, expansion and differentiation of B cells (Ansel et al., 1999; Breitfeld et al., 2000). Defects in germinal center (GC) formation and antibody production is associated with the deficiency in CD4^+^ T cell derived signals like CD40L-CD40 interaction and IL-21 cytokine (Allen et al., 1993; Tangye and Ma, 2020). Clearly, a synchronized and robust cognate interaction between CD4^+^ T-cell and B cell is the basis of a productive humoral output in terms of memory B cells, plasma cells and antibody production. Thus, the extent of help stimulus provided by CD4^+^ T cells may correlate with the magnitude and quality of humoral response generated after infection or vaccination.

The *ex vivo* T-B co-culture assay is the attempt to recapitulate the physiological process of T-B cognate interactions to measure the ability of CD4^+^ T-cell subsets in providing help to B cells. The most commonly used *ex-vivo* method involves co-culturing of CD4^+^ T cells with B cells in presence of super-antigen like Staphylococcus enterotoxin B (SEB) for 7-12 days. The importance of this method is that it allows cross-linking between CD4^+^ T cells and B cells through TCR-pMHC-II interactions, thus resembling the physiological condition of T cells engagement with B cells. This method has been widely used to compare the helper potential of CXCR5^+^ Tfh subsets or the CXCR5^-^ non-Tfh subsets expressing activation markers in various infections and autoimmune diseases (Bentebibel et al., 2013; Caielli et al., 2019; Locci et al., 2013; Morita et al., 2011; Rao et al., 2017). Indeed, superantigen based co-culture method was instrumental in revealing the potential and superiority of Tfh subsets in providing help to B cells. However, the superantigen based method do not signify the antigen-specific cognate interactions between T cells and B cells, and thus cannot be utilized for defining the help potential of CD4^+^ T cell subset in antigen-specific manner. Recently, MHCII-restricted (CD4^+^) peptides are used to determine the quantity of antigen-specific CD4^+^ T cells in the PBMCs (Reiss et al., 2017). The stimulation of T-B cocultures using peptides was helpful in defining the B cell helper function of HIV-specific CD4^+^ T cells (Del Alcazar et al., 2019). However, the use of antigen-specific T-B co-cultures is limited may be due to the inconsistent detection of plasma cell output and the poor sensitivity in examining other immunologic parameters of the productive T-B cross-talk.

In fact, antigen-specific T-B co-culture utilizes the function of B cells both as the antigen presenting cells (APCs), in order to activate the CD4^+^ T cells, as well as the recipient of helper signals from the activated cognate CD4^+^ T cells. Thus, the efficiency and success of T-B co-cultures completely dependent on the ability of B cells to prime and activate antigen-specific CD4^+^ T cells. Further, the survivability of B cells in the *ex vivo* cultures is also crucial, because these cells cannot survive longer in absence of optimal survival signals (Kraus et al., 2004; Schweighoffer et al., 2013). In the peptide-stimulated co-cultures, it’s unlikely that B cells are efficiently performing its APCs function or getting sufficient signals for its optimal survival in absence of BCR-mediated signaling. This could also be a probable reason for poor sensitivity of the conventional T-B co-cultures in producing the consistent antigen-specific responses.

The current pandemic signifies the need of multiple immunologic parameters to be measured while establishing the correlates of protection from COVID-19 vaccine or while determining the traits of broad protective immunity in natural infection. The CD4^+^ T cells are widely implicated in the less severe outcome of SARS-CoV-2 infection (Braun et al., 2020; Rydyznski Moderbacher et al., 2020; Tan et al., 2021). Indeed, efficient Tfh responses and the ensuing enhanced plasmablasts frequency and antibody production are associated with mild outcome of COVID-19 (Kuri-Cervantes et al., 2020). Because the T-cell help is indispensable for good quality B cell and antibody responses, the qualitative assessment of the helper function of memory CD4^+^ T cells may serve as an immunologic parameter for defining functional memory responses. Here, we describe an antigen-specific T-cell and B cell co-culture assay that is highly consistent in measuring the help potential of memory CD4^+^ T cells. We investigated the role of monocytes that are the abundantly available circulating myeloid cells in enhancing the efficiency of autologous T-B co-cultures for antigen-specific analysis. Using *M. tuberculosis* (Mtb) specific settings, we demonstrate that monocytes are superior antigen presenting cells (APCs) than B cells in the early stages of co-cultures to prime and activate antigen-specific CD4^+^ T cells. Moreover, in addition to functioning as APCs, monocytes promote the survivability of B cells in cultures mainly by delivering essential growth factors like SCGF/CLEC11A and BAFF. We found SCGF/CLEC11A (stem cell growth factor or C-type lectin domain family 11 member A) as a new class of growth factor that is essential for the B cell survival. We further demonstrate the application of highly efficient monocyte supplemented T-B co-cultures in assessing the helper function of SARS-CoV-2 spike specific CD4^+^ T cells in the COVID-19 recovered donors. The assessment was performed by quantifying the plasma cell differentiation, antibody production, antibody secreting cells and IL-21 secreting T cells in the SARS-CoV-2 specific co-cultures. The method described here is highly sensitive, quantitative, yields the output that is substantiated by several immunologic parameters of T-dependent B cell responses, and is applicable to assess the B cell help function of SARS-CoV-2-specific memory CD4^+^ T cells.

## Results

### Survival of B cells is enhanced by supplementing T-B co-cultures with the monocytes

The availability of functionally viable cells is important for autologous T and B co-cultures. Thus, to examine the survivability of T and B cells in the autologous co-cultures, we sorted the cells from PBMCs of healthy blood donors (**Figure S1**) and co-cultured 60,000 cells of memory CD4^+^ T cells and CD20^+^ B cells in 1:1 ratio for various days up to 12 days, without any exogenous antigen. As expected, in absence of antigen-specific stimulation, total cells did not survive in the culture and only 3.8±1.3% (mean±sem) total cells survived till 12 days (**Figure S2 A-B**). Number of live B cells was significantly reduced after 12 days of co-culture as compared with 9 days (**Figure S2 C**; 9 days: 843±245; 12 days: 93±25; P=0.02). Both naïve and memory B cells died gradually till 12 days of co-culture (**Figure S2 D**). Similarly, substantial decline in number of live T cells was observed in 12 days as compared with 9 days (**Figure S2 E**; 9 days: 2341±613; 12 days: 358±120; P=0.008). *Mycobacterium tuberculosis* (Mtb) specific CD4^+^ T cells are present in majority of the healthy blood donors in our cohort (Unpublished data). Thus, for antigen specific stimulations, we used Mtb specific pool of CD4^+^ T-cell peptides (Mtb peptides). In all the time points analyzed, no significant variation in B cell survivability was observed in presence of Mtb peptides over the unstimulated conditions (**Figure S3 A**). However, in Mtb peptides stimulated cultures, a significant increase in live T-cell count was observed at Day 9 over the unstimulated condition (**Figure S3 B**; unstimulated: 3473±1382; Mtb peptides: 10490±3022; P=0.03). Like total B cells, no significant increase was observed in number of plasma cells (CD20^lo^CD38^hi^) in presence of Mtb peptides as compared to unstimulated condition (**Figure S3 C-D**). We then quantified the activated CD4^+^ T cells using CD38 and PD-1 and found that a significant number of CD38^+^PD-1^+^CD4^+^ T cells was detected as early as 3 days, which peaked at Day 9 (**Figure S3 E-F**; 9 days: unstimulated, 58±24; Mtb peptides, 3948±1076; P=0.001). Therefore, with the maximal activation of T cells without significant loss in the viability of both T and B cells, we concluded 9 days of co-culture as the optimal duration for further experiments.

The myeloid cells are implicated in supporting the survivability of lymphoid cells, mainly B cells (Epron et al., 2012; Mueller et al., 2007). Thus, to enhance the survivability of cells in co-cultures, we attempted supplementing the cultures with monocytes that are abundantly available blood myeloid cells. To determine the impact of monocytes on survivability of B cells in the *ex vivo* cultures, we cultured 60,000 CD20^+^ B cells with CD14^+^ monocytes (M) for 9 days in various M to B ratios, 1:4, 1:2 and 1:1 in absence of any exogenous antigen. Titration of monocytes to B cells revealed that survivability of B cells in presence of monocytes increased significantly at 1:2 ratio compared to B cells alone (**Figure S4 C**; 1:2 ratio vs B cells alone; P=0.0002), however the presence of monocytes in 1:2 and 1:1 ratio had the comparable impact on B cell survivability (**Figure S4 C**; 1:2 M to B ratio: 1183±269; 1:1 M to B ratio: 1543±233; ns). Thus, we used the monocytes to B cell ratio of 1:2 for further experiments. We then compared the survivability of B cells and T cells in the cultures with or without monocytes, in the unstimulated autologous cultures. We found that count of live B cells in presence of monocytes (B+M) was significantly higher as compared to B cells alone (**Figure 1 B-C**; B alone: 110±43; B+M: 1644±245; P<0.0001), whereas survivability of B cells was not significantly different over B cells alone when cultured with T cells (T+B) (**Figure 1C**; T+B: 501±77; P=0.1). Monocytes enhanced the survivability of B cells by 30-fold (median), in contrast to T cells that showed only 9-fold increase in B cells survivability (**Figure 1D**; B+M versus B+T: P=0.009). Moreover, monocytes enhanced the survivability of both naive (CD27^-^ B cells) and memory (CD27^+^ B cells) compartments, with similar impact on both the compartments, in comparison with B+T (**Figure 1E**; Naïve: B+M: 68-fold; B+T: 13-fold; P=0.02; Memory: B+M: 24-fold; B+T: 8-fold; P=0.04). Unlike B cells, T cells survived more efficiently when co-cultured with monocytes or B cells, as compared to alone culture (**Figure 1 F-G**; T alone, 2789±330; T+M: 6122±663; P<0.0001; and T+B: 4574±463; P=0.005). Survivability of T cells was comparable in presence of either monocytes or B cells (**Figure 1H**; T+M: 2.1-fold; T+B: 1.7-fold; P=0.15). Altogether, the data suggest that monocytes supplementation promotes the survivability of both T and B cells in autologous cultures, with a substantially higher efficiency of increasing the survivability of B cells.

**Figure 1.**
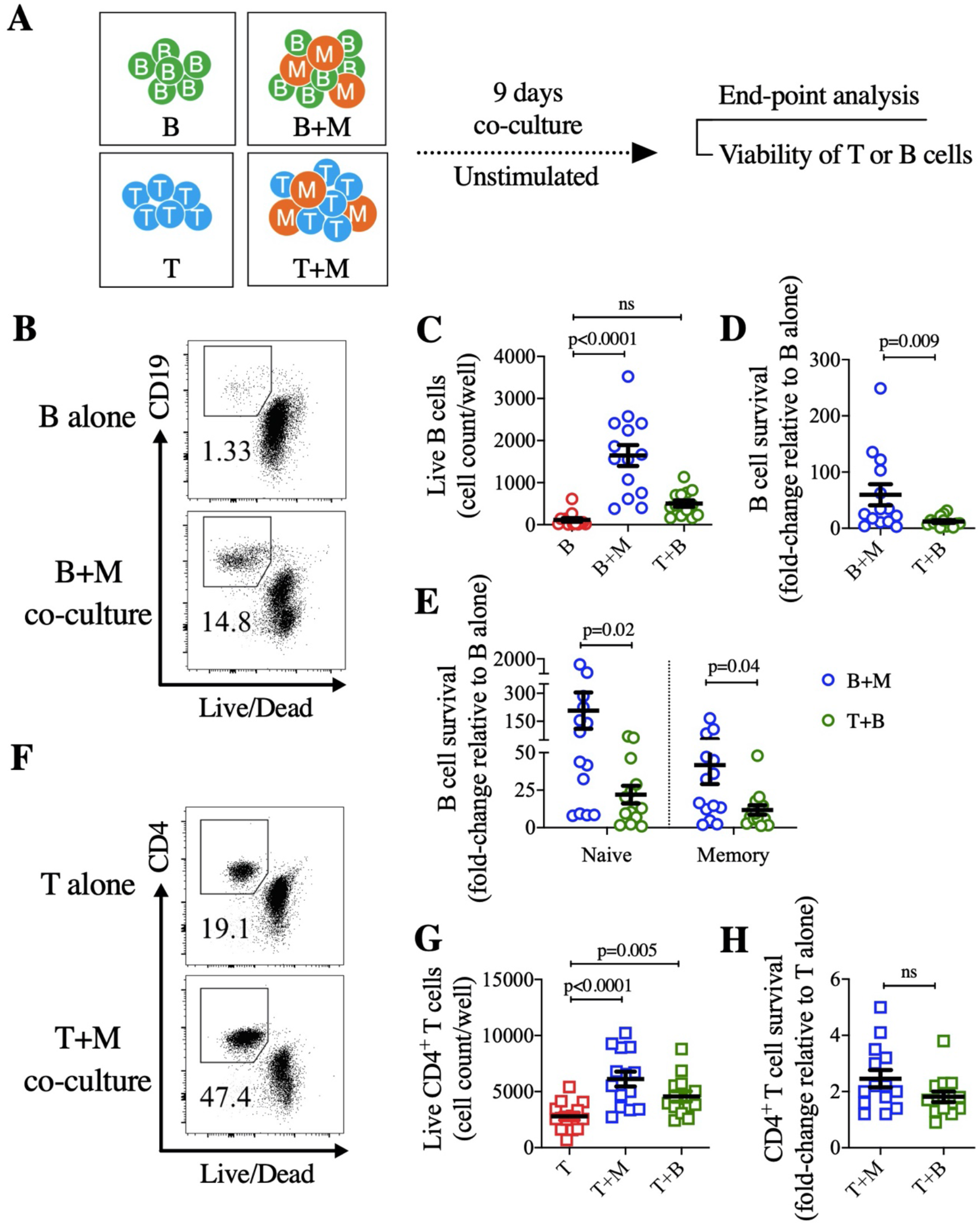
Enhanced survivability of B cells in presence of monocytes in *ex vivo* co-cultures. (**A)** Schematic representation for examining the impact of monocytes on survival of B cells and T cells in *ex vivo* co-culture, in absence of any exogenous antigen (unstimulated). (**B)** Representative dot plots for the flow cytometric analysis of live B cells (live CD19^+^ cells) in absence (B alone) or presence of monocytes (B+M co-culture). (**C)** Count of live B cells shown in co-cultures of B cells alone (B), B cells + monocytes (B+M) and B cells + T cells (T+B). (**D)** Extent of B cell survival in B cells + monocytes (B+M) and B cells + T cells (T+B) co-culture. The fold-change in survival was calculated by dividing the count of live B cells in B+M or B+T co-culture with the count in B cell alone culture. (**E)** Magnitude of B cell survivability in naïve (CD27^-^ B cells) and memory (CD27^+^ B cells) compartments in either B+M (blue square) or B+T (green triangle) co-cultures. The fold-change in survival was calculated as explained in panel D. (**F)** Representative dot plots for the flow cytometric analysis of live CD4^+^ T cells (live CD4^+^ cells) in absence (T alone) or presence of monocytes (T+M co-culture). (**G)** Count of live CD4^+^ T cells shown in T alone, T+M and T+B co-cultures. (**H)** Extent of T cell survival in T+M or T+B co-culture. The fold-change in T-cell survival was calculated by dividing the count of live CD4^+^ T cells in T+M or T+B co-culture with the count in T alone culture. Data is represented as mean ± sem, with each dot representing one donor. Data represent the pool of three independent experiments. Statistical comparisons were performed by (C and G) one-way ANOVA followed by Bonferroni’s multiple comparisons test, and (D, E and H) two-tailed Mann-Whitney test. ns: non-significant.

### Monocytes support in an efficient qualitative assessment of antigen-specific helper T cells

The quantitative measurement of the help provided by CD4^+^ T cells to the B cells for differentiation into plasma cells and antibody secretion is a benchmark parameter to determine the quality of CD4^+^ T cells. Thus, we utilized the monocytes supplementation in T-B co-culture assays to address whether adding monocytes to T-B co-cultures only enhances the survivability of cells or it also augments the productivity of T-B cognate interactions in the antigenic stimulations. We co-cultured 60,000 cells of memory CD4^+^ T cells and CD20^+^ B cells in absence or presence of 30,000 cells of CD14^+^ monocytes for 9 days (**Fig. 2A**). Absence of any exogenous antigen (unstimulated) condition was used as a control for the antigen non-specific background response. For antigen-specific stimulation, we used the *M. tuberculosis* whole cell lysate (Mtb Lysate) and above used Mtb peptides. Mtb lysate is considered as the complex mixture of various *M. tuberculosis* antigens including proteins, lipids and carbohydrates present in the bacterial cell (Brennan, 2003; Jankute et al., 2015) and therefore, they need to be internalized and processed by monocytes or B cells before presentation to antigen-specific T cells. On contrary, *M. tuberculosis* specific pool of CD4^+^ T-cell peptides (Mtb peptides) does not need processing and can quickly be presented by monocytes or B cells. In T-B co-culture, the plasma cell count was not significantly different between the Mtb peptide stimulation and the unstimulated condition (**Figure 2B, C**; T+B; Unstimulated: 11±4; Mtb peptides: 155±89; ns). However, stimulation with Mtb lysate was able to induce significantly higher plasma cells over the unstimulated control (**Figure 2B, D**; T+B; Unstimulated: 108±45; Mtb lysate: 760±227; P=0.03). Supplementation of monocytes to T-B co-cultures significantly enhanced the output of plasma cells in presence of both the antigens, Mtb peptides (**Figure 2B, C**; T+B+M; Unstimulated: 17±7; Mtb peptides: 1455±396; P=0.006) and Mtb lysate (**Figure 2B, D**; T+B+M; Unstimulated: 281±95; Mtb lysate: 3004±926; P=0.01). In the B+M cultures, in absence of T cells, no significant number of plasma cells was detected in unstimulated and stimulated conditions with any of the antigens (**Figure 2 B-D)**. When we compared the background (unstimulated) subtracted number of plasma cells, we found monocytes supplemented T-B co-cultures (T+B+M) were highly efficient in plasma cell output compared with T-B co-cultures (T+B) in presence of both the antigens, Mtb peptides (**Figure 2E**; T+B versus T+B+M; P=0.005) and Mtb lysate (**Figure 2F**; T+B versus T+B+M; P=0.03). We further confirmed this observation by quantifying the level of secreted immunoglobulin IgG in supernatant and found that secreted IgG level in T+B+M was superior than the IgG level in T+B, in case of both the antigens, Mtb peptides (**Figure 2G**; T+B, 493±251 ng/mL; T+B+M, 3454±689 ng/mL; P=0.004) and Mtb lysate (**Figure 2H**; T+B, 1268±324 ng/mL; T+B+M, 3507±786 ng/mL; P=0.008). We then quantified the activated CD4^+^ T cells in the co-cultures by using the ICOS and PD-1 co-expression on CD4^+^ T cells. Number of ICOS^+^PD-1^+^CD4^+^ T cells was detected significantly in all the co-cultures (T+M, T+B, T+B+M) in presence of both of the antigens, Mtb peptides and Mtb lysate when compared with respective unstimulated controls (**Figure 2 I-K**). Similar to plasma cell differentiation, monocytes supplementation to T-B co-cultures (T+B+M) enhanced the magnitude of ICOS^+^PD-1^+^CD4^+^ T cells compared with T-B co-cultures (T+B) in presence of Mtb peptides (**Figure 2L**; T+B, 1904±635; T+B+M, 5983±1,263; P=0.001) and Mtb lysate (**Figure 2M**; T+B, 2318±714; T+B+M, 5247±1,509; P=0.05). Hence, the data from autologous antigen-specific cultures using the Mtb antigens clearly suggest that monocyte supplementation to T-B co-cultures increases the productivity of cognate T-B interactions. Moreover, it also enhances the assay sensitivity in detecting the plasma cell output and T-cell activation.

**Figure 2.**
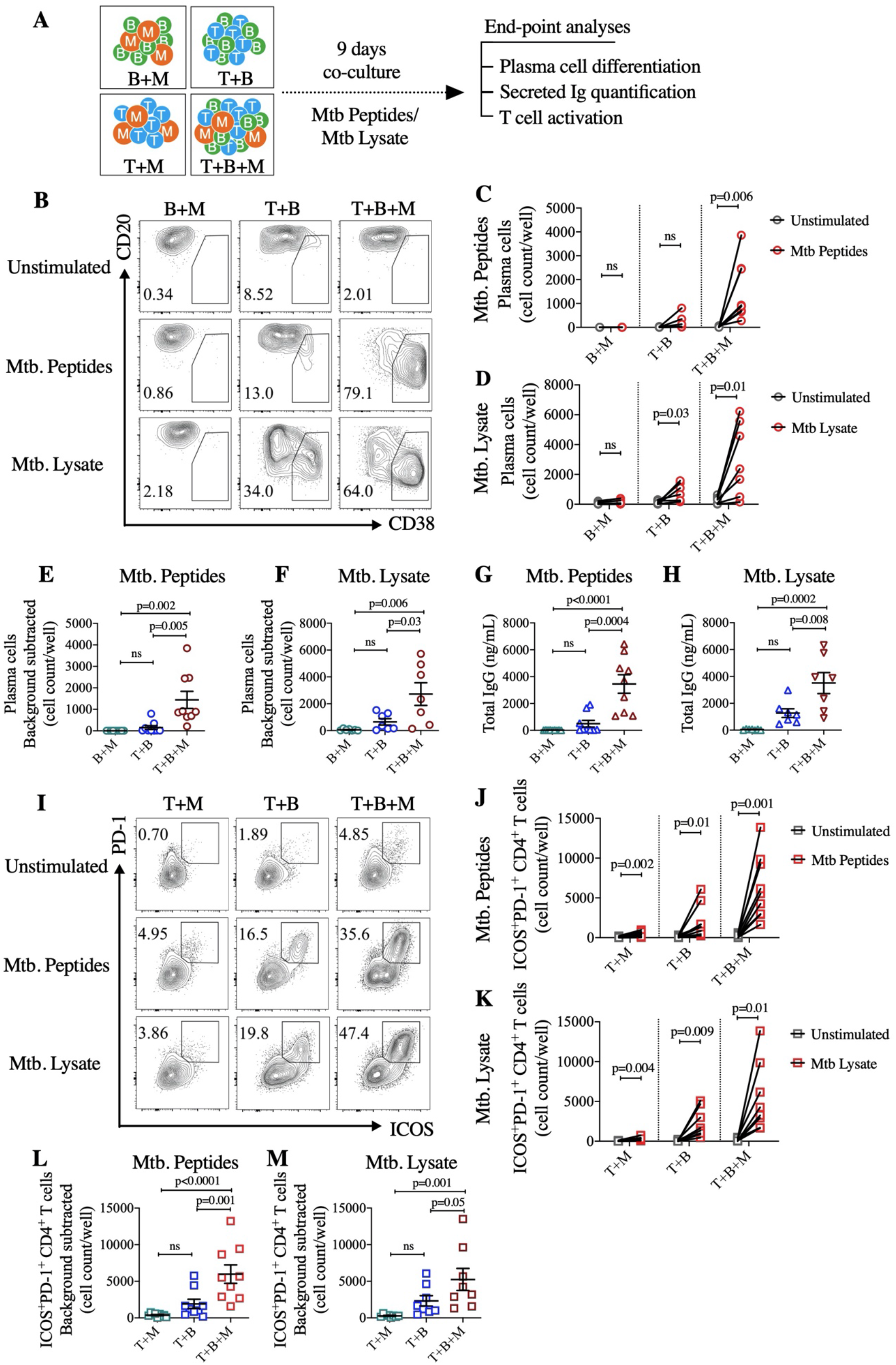
Monocytes in the T-B co-cultures support an efficient qualitative assessment of the helper potential of *M. tuberculosis* specific memory CD4^+^ T cells. **(A**) Schematic representation for the analysis of helper function of *M. tuberculosis* specific memory CD4^+^ T cells from healthy individuals in the T-B co-culture in presence of autologous monocytes. **(B)** Representative contour plots for the flow cytometric analysis of plasma cells (CD20^lo^CD38^hi^cells gated in CD27^+^ B cells) in B cells + Monocytes (B+M), T cells + B cells (T+B) and T cells + B cells + Monocytes (T+B+M) co-cultures in no stimulation control condition (unstimulated) and in stimulated conditions either with *M. tuberculosis* (Mtb.) peptides or Mtb whole cell lysate (Mtb. Lysate). Plasma cell count shown in B+M, T+B and T+B+M co-cultures in stimulation with (**C**) Mtb. peptides and (**D**) Mtb. whole cell lysate, in comparison with their unstimulated control conditions. Line connecting grey and red open circles represent the same donor in the unstimulated and stimulated conditions, respectively. The plasma cell count in unstimulated control condition was subtracted as the background from the plasma cell count in stimulated conditions. Shown is the background subtracted plasma cell count in stimulated conditions with (**E**) Mtb. peptides and (**F**) Mtb. whole cell lysate. Total immunoglobulin G (IgG) was quantified in the supernatants of B+M, T+B and T+B+M co-cultures in (**G**) Mtb. peptides stimulated condition and (**H**) Mtb. lysate stimulated condition. **(I)** Representative contour plots for the flow cytometric analysis of ICOS^+^PD-1^+^CD4^+^ T cells (gated in CD4^+^T cells) in T+M, T+B and T+B+M co-cultures in unstimulated control condition and stimulated conditions either with *M. tuberculosis* (Mtb.) peptides or Mtb. whole cell lysate (Mtb. Lysate). ICOS^+^PD-1^+^CD4^+^ T cell count shown in T+M, T+B and T+B+M co-cultures in stimulated conditions (red circle) with (**J**) Mtb. peptides and (**K**) Mtb. lysate, in comparison with their unstimulated control conditions (grey circle). Background subtracted ICOS^+^PD-1^+^CD4^+^ T cell count in (**L**) Mtb peptides stimulated condition and (**M**) Mtb. whole cell lysate stimulated condition, calculated as ICOS^+^PD-1^+^CD4^+^ T cells in stimulated condition minus ICOS^+^PD-1^+^CD4^+^ T cells in unstimulated control condition. Data is represented as mean ± sem, with each dot representing one donor. Data represent the pool of two independent experiments. Statistical comparisons were performed by (C-D and J-K) two-tailed paired t test and (E-H and L-M) one-way ANOVA followed by Bonferroni’s multiple comparisons test. ns: non-significant.

### Qualitative evaluation of helper functions of SARS-CoV-2 specific memory CD4^+^ T cells using monocyte supplemented T-B co-cultures

Several studies have highlighted the crucial role of CD4^+^ T cells in protective response to SARS-CoV-2. Others and we have shown that immune memory in terms of CD4^+^ T cells and B cells is persistent after several months in COVID-19 recovered subjects (Ansari et al., 2021; Cohen et al., 2021; Dan et al., 2021; Rodda et al., 2021). However, the helper quality of memory CD4^+^ T cells in diverse outcome of COVID-19 or in response to different vaccine preparations is not studied. There is an urgent need of sensitive assays that can help in attempting the qualitative evaluation of SARS-CoV-2 specific CD4^+^ T cells. Thus, we investigated the monocyte supplemented T-B co-culture assay in evaluating the helper functions of SARS-CoV-2 specific memory CD4^+^ T cells in COVID-19 recovered subjects. We co-cultured 60,000 memory CD4^+^ T cells and CD20^+^ B cells in absence or presence of 30,000 CD14^+^ monocytes for 9 days (**Figure 3A**). Unstimulated condition was used as a control for the antigen non-specific background response. For antigen-specific stimulation, we used the SARS-CoV-2 spike peptides (CoV-S peptides) and SARS-CoV-2 full length spike protein (CoV-S protein). We detected a higher number of plasma cells in T-B co-culture (T+B) in presence of CoV-S peptides pool (**Figure 3B, C**; T+B; Unstimulated: 23±11; CoV-S peptides: 1087±252; P=0.003), but not in presence of CoV-S protein in comparison with the unstimulated controls (**Figure 3B, D**; T+B; Unstimulated: 11±4; CoV-S protein: 75±37; ns). However, monocyte supplemented T-B co-culture (T+B+M) allowed the efficient detection of significantly higher output of plasma cells in the presence of CoV-S peptides (**Figure 3B, C**; T+B+M; Unstimulated: 41±16; CoV-S peptides: 2840±553; P=0.001) as well as in the presence of CoV-S protein (**Figure 3B, D**; T+B+M; Unstimulated: 53±17; CoV-S protein: 1244±398; P=0.01). Comparing the background subtracted count of plasma cells revealed that monocyte supplemented T-B co-culture (T+B+M) significantly enhanced the magnitude of plasma cell differentiation compared with non-supplemented T-B co-culture (T+B) in presence of both the antigens, CoV-S peptides (**Figure 3E**; T+B versus T+B+M; P=0.02) and CoV-S protein (**Figure 3F**; T+B versus T+B+M; P=0.01). The higher IgG levels in response to both the antigens, CoV-S peptides (**Figure 3G**; T+B vs T+B+M; P=0.01) and CoV-S protein (**Figure 3H**; T+B vs T+B+M; p=0.01), further substantiated the superior plasma cell output in monocyte supplemented conditions. Monocyte supplementation to T-B co-culture also enhanced the antigen-specific IgG levels as the secreted CoV-S protein specific IgG level in T+B+M was significantly ∼20-fold higher in magnitude than the IgG level in T+B co-cultures (**Figure 3I**; T+B vs T+B+M; P=0.01). The quantitative analyses of antibody secreting cells (ASCs) (**Figure 3 J-K**) corroborated with our findings of higher plasma cell output and IgG production, as total IgM and IgG ASCs were increased in monocyte supplemented T-B co-cultures (T+B+M) as compared with monocyte non-supplemented T-B co-culture (T+B) (**Figure 3L;** IgM ASCs; T+B vs T+B+M; P=0.0009; IgG ASCs; T+B vs T+B+M; P=0.002). Similarly, CoV-S specific IgM and IgG ASCs were also enhanced in presence of monocytes (T+B+M) by 4 and 5-fold, respectively, over the T+B co-cultures alone (**Figure 3M;** IgM ASCs; T+B vs T+B+M; P=0.005; IgG ASCs; T+B vs T+B+M; P=0.03). Altogether, the monocyte supplemented T-B co-culture was superior over the T-B alone co-culture in detecting the T-dependent plasma cell differentiation and antibody generation in response to SARS-CoV-2 virus antigens.

**Figure 3.**
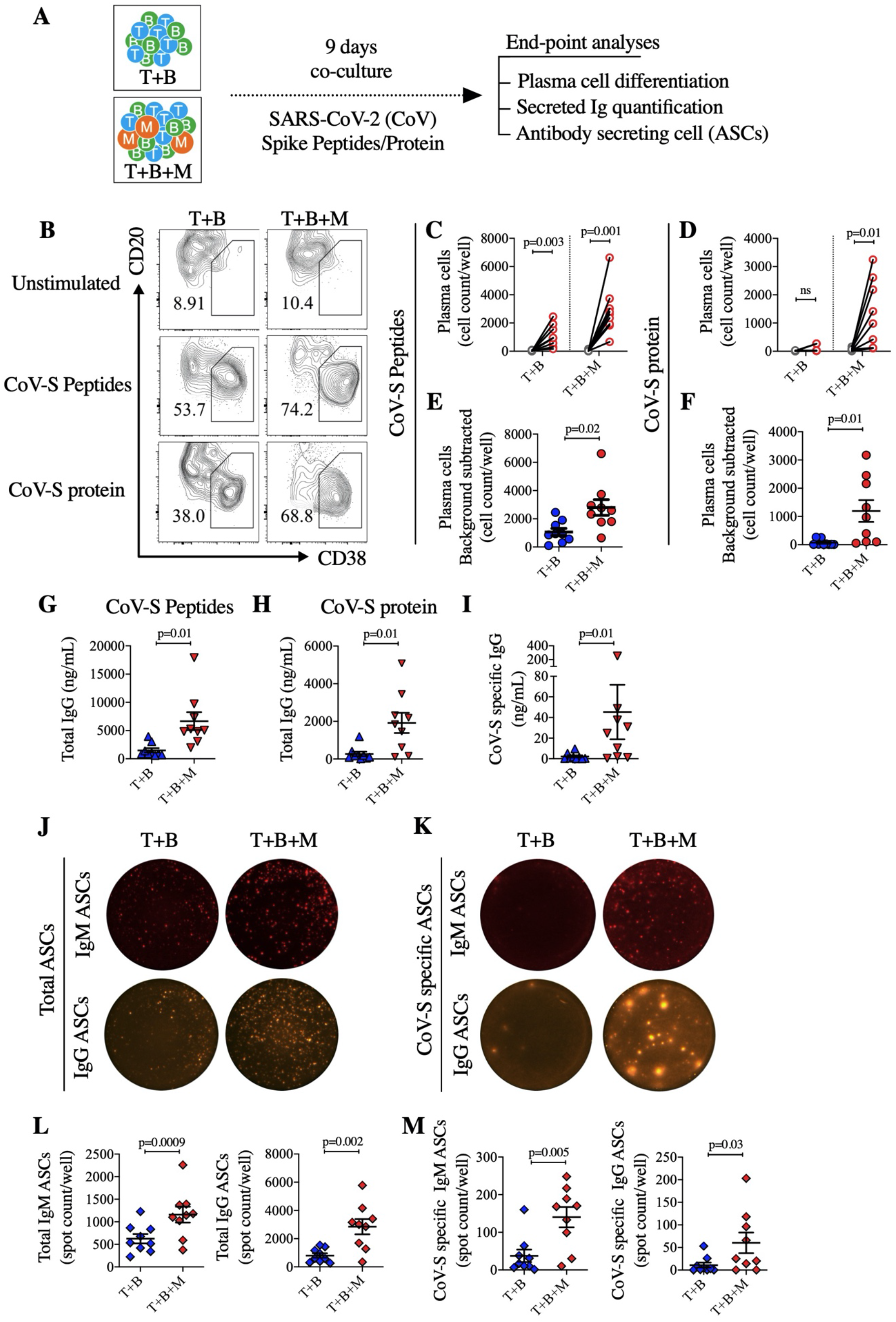
Monocyte supplemented T-B co-culture assay permits the qualitative evaluation of helper function of SARS-CoV-2 specific memory CD4^+^ T cells in COVID-19 recovered subjects. **(A**) Schematic representation for the evaluation of helper function of SARS-CoV-2 specific memory CD4^+^ T cells in T-B co-culture assay in presence of monocytes. **(B)** Representative contour plots for the flow cytometric analysis of plasma cells (CD20^lo^CD38^hi^cells gated in CD27^+^ B cells) in T+B and T+B+M co-cultures in unstimulated control condition or stimulated conditions with SARS-CoV-2 spike peptides (CoV-S peptides) and SARS-CoV-2 full length spike protein (CoV-S protein). Shown is the Plasma cell count in T+B and T+B+M co-cultures in (**C**) CoV-S peptides stimulated condition and (**D**) CoV-S protein stimulated condition, in comparison with their unstimulated control conditions. Line connecting grey and red open circles represent the same individual in the unstimulated and stimulated conditions, respectively. Background subtracted plasma cell count in (**E**) CoV-S peptides stimulated condition and (**F**) CoV-S protein stimulated condition, calculated as plasma cells count in stimulated condition minus the count in unstimulated control condition. Total immunoglobulin G (IgG) was quantified in the supernatants of T+B and T+B+M co-cultures in (**G**) CoV-S peptides stimulated condition and (**H**) CoV-S protein stimulated condition. (**I**) SARS-CoV-2 spike protein (CoV-S protein) specific IgG was quantified in the supernatants of T+B and T+B+M co-cultures in CoV-S peptides stimulated condition. Representative FluoroSpot images for the **(J)** total IgM and IgG antibody secreting cells (ASCs) and **(K)** the SARS-CoV-2 spike protein (CoV-S protein) specific IgM- and IgG-ASCs, measured in the T+B and T+B+M co-cultures after 9 days of stimulation with SARS-CoV-2 spike peptides. Shown are the spot count per well in T+B and T+B+M co-cultures in CoV-S peptides stimulated conditions in (**L**) Total IgM ASCs (*left*) and total IgG ASCs (*right*) and (**M**) CoV-S specific IgM ASCs (*left*) and CoV-S specific IgG ASCs (*right*). Data is represented as mean ± sem, with each dot representing one donor. Data represent the pool of two independent experiments. Statistical comparisons were performed by (C and E) multiple t tests corrected using Holm-Sidak method, (C-H and L-M) two-tailed paired t test (I) two-tailed Wilcoxon matched-pairs signed rank test. ns: non-significant.

We next examined the level of activated CD4^+^ T cells in these monocyte supplemented T-B co-cultures. In absence of monocyte supplementation to T-B co-cultures (T+B), a significant number of ICOS^+^PD-1^+^CD4^+^ T cells were detected in presence of CoV-S peptides (**Figure 4B, C**; T+B; Unstimulated: 576±211; CoV-S peptides: 5316±1,121; P=0.001), but not in the presence of CoV-S protein (**Figure 4B, D**; T+B; Unstimulated: 418±137; CoV-S protein: 2061±923; ns), in comparison with the unstimulated controls. The monocyte supplementation (T+B+M) showed significantly higher number of ICOS^+^PD-1^+^CD4^+^ T cells in presence of both of the antigens, CoV-S peptides (**Figure 4B, C**; T+B+M; Unstimulated: 2511±745; CoV-S peptides: 18122±3,310; P=0.0004) and CoV-S protein (**Figure 4B, D**; T+B+M; Unstimulated, 2630±797; CoV-S protein, 10640±3104; p=0.02). Like plasma cell output, consistently higher magnitude of ICOS^+^PD-1^+^ activated CD4^+^ T cells was observed in presence of both of the antigens, CoV-S peptides (**Figure 4E**; T+B versus T+B+M; P=0.001) and CoV-S protein (**Figure 4F**; T+B versus T+B+M; P=0.04). The IL-21 cytokine secreted by CD4^+^ T cells is an important factor for the T-cell dependent plasma cell differentiation (Bryant et al., 2007; Ettinger et al., 2005; Kuchen et al., 2007). Thus, we compared the magnitude of IL-21 secreting cells in monocyte supplemented or non-supplemented T-B co-cultures, after restimulation with CoV-S peptides (**Figure 4G**). As expected, the number of IL-21 secreting cells (spot forming cells, SFCs) was significantly ∼4-fold higher in T+B+M co-cultures over the T+B alone cultures (**Figure 4H;** T+B: 48±14; T+B+M: 178±41; P=0.006). We further quantified several secreted soluble factors in the monocyte supplemented or non-supplemented T-B co-culture in presence of CoV-S protein stimulation. We detected significantly higher levels of various cytokines in monocyte supplemented conditions like IL-2, IFN-γ, IL-4, IL-17, sCD40L, IL-10 and IL15 (**Figure 4I**). Majority of these cytokines may help in the B cell proliferation, plasma cell differentiation and isotype switching (Vazquez et al., 2015). However, the secreted IL-21 was below the limit of quantification and was not detectable in the cytokine multiplex assay. In presence of monocytes (T+B+M), we also detected highly up-regulated levels of pro-inflammatory cytokines such as IL-1β, TNF-α and IL-6 (**Figure 4I**). Thus, we confirmed that monocyte supplementation to antigen specific T-B co-cultures enables the activation of functionally potent SARS-CoV-2 specific memory CD4^+^ T cells, which deliver the appropriate help signals to B cells. Altogether, the results clearly suggest that the monocyte supplementation to the T-B co-culture enhances the sensitivity for efficient qualitative assessment of the SARS-CoV-2 specific memory CD4^+^ T cells.

**Figure 4.**
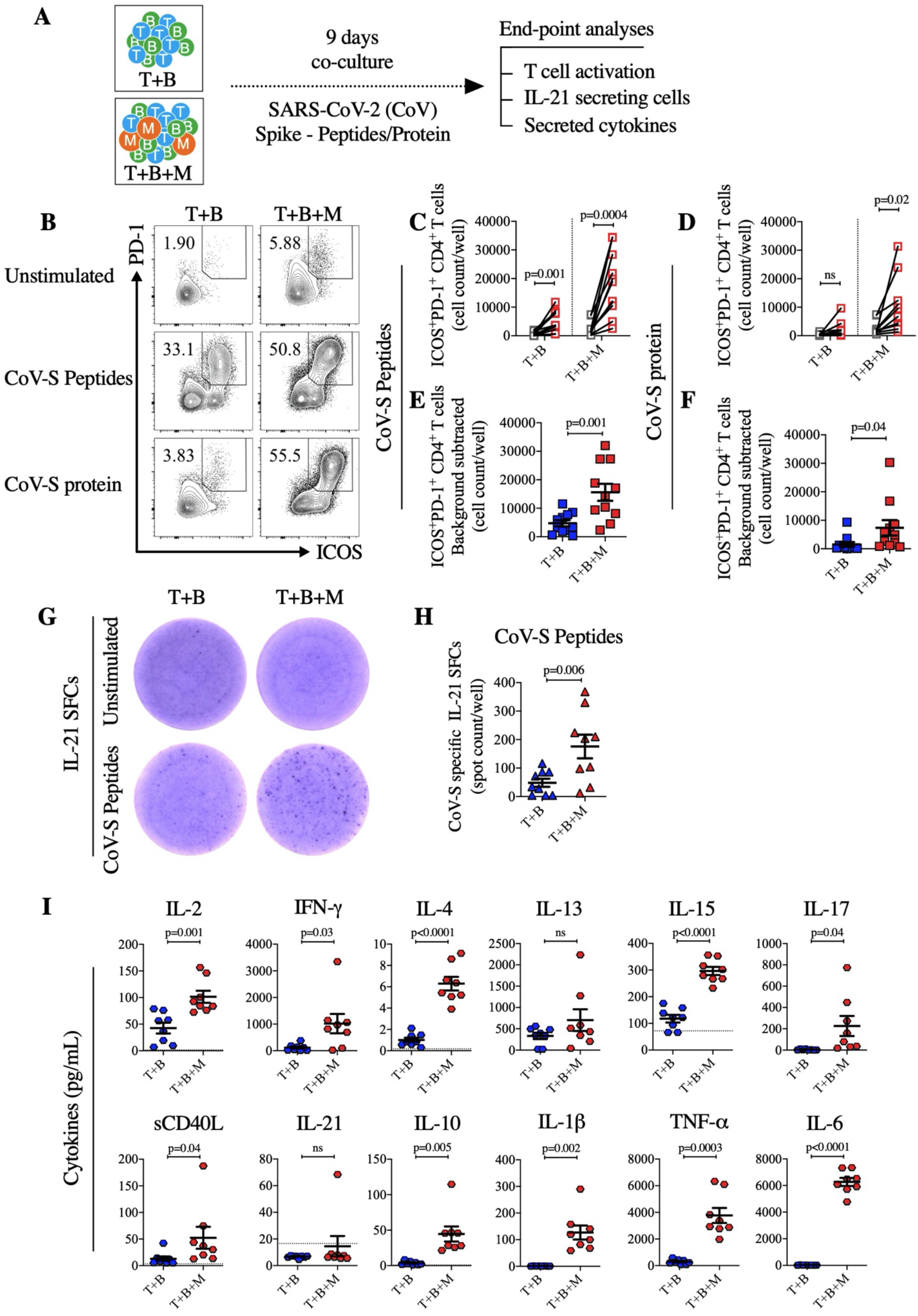
Enhanced activation and functional potential of SARS-CoV-2 specific memory CD4^+^ T cells in monocytes supplemented T-B co-cultures. **(A**) Schematic representation for the analysis of SARS-CoV-2 specific memory CD4^+^ T cells in T-B co-culture in presence of monocytes. **(B)** Representative contour plots for the flow cytometric analysis of ICOS^+^PD-1^+^CD4^+^ T cells (gated in CD4^+^ T cells) in T+B and T+B+M co-cultures in unstimulated control condition or stimulated conditions with SARS-CoV-2 spike peptides (CoV-S peptides) and SARS-CoV-2 spike protein (CoV-S protein). ICOS^+^PD-1^+^CD4^+^ T cell count shown in T+B and T+B+M co-cultures in (**C**) CoV-S peptides stimulated condition and (**D**) CoV-S protein stimulated condition, in comparison with their unstimulated control conditions. Line connecting grey and red open circles represent the same individual in the unstimulated and stimulated conditions, respectively. Background subtracted ICOS^+^PD-1^+^CD4^+^ T cell count in (**E**) CoV-S peptides stimulated condition and (**F**) CoV-S protein stimulated condition, calculated as ICOS^+^PD-1^+^CD4^+^ T cells count in stimulated condition minus the count in unstimulated control condition. **(G)** Representative ELIspot well images for the IL-21 spot forming cells (SFCs) detected in the CD4^+^ T cells either unstimulated or re-stimulated with CoV-S peptides after 9 days of T+B and T+B+M co-cultures. (**H**) CoV-S specific IL-21 SFCs shown as spot count per well in T+B and T+B+M co-cultures in the CoV-S peptides stimulated condition. (**I**) Quantification of secreted cytokines in supernatant of T+B and T+B+M co-cultures in the CoV-S protein stimulated condition. Dotted horizontal line represents the lowest limit of quantification (LLOQ). Data is represented as mean ± sem, with each dot representing one donor. Data represent the pool of two independent experiments. Statistical comparisons were performed by (C-F and H-I) two-tailed paired t test. ns: non-significant.

### Monocytes act as superior antigen presenting cells for initial priming and activation of antigen-specific memory CD4^+^ T cells

Monocytes played a vital role in enhancing the outputs of antigen-specific T-B co-cultures by promoting efficient T cell activation and plasma cell differentiation. We thus examined why adding monocytes enhanced the activation of CD4^+^ T cells and augmented the productivity of T-B cognate interactions, while B cells alone was not sufficient. Among the classical antigen presenting cells (APCs), CD14^+^ monocytes in healthy donor’s PBMCs are present in the proportion of 14-23% (**Figure S5 A-B;** 18±1%) which is almost double than the frequency of CD20^+^ B cells (**Figure S5 A-B;** 9-11%). CD14^+^ monocytes represent relatively more homogenous population compared with CD20^+^ B cells, which is quite heterogenous comprising approximately 70% of naive, 10% of unswitched and 14% of switched memory cells (**Figure S5 A-B**). Though the monocytes and all the subsets in CD20^+^ B cells constitutively express similar level of HLA-DR, these APCs differ in the expression of co-stimulatory molecules, CD80 and CD86 (**Figure S5 C-D**). Nearly all the monocytes constitutively express CD86 but not CD80 whereas none of B cell subsets express CD86 (**Figure S5 C-D**). Roughly a half (55±3%, data not shown) of the memory B cells expresses CD80. Thus, CD20^+^ B cells have less potential to provide costimulatory signals than CD14^+^ monocytes during the initial priming of memory T cells. To determine which one of monocytes or B cells is superior in the initial priming and activation of memory T cells during the early stage of co-culture, we analyzed the co-cultures of 60,000 memory CD4^+^ T cells in presence of either 30,000 CD14^+^ monocytes (T+M) or 60,000 CD20^+^ B cells (T+B) or combination of both (T+B+M), in the presence of Mtb antigens (Mtb peptides and Mtb lysate) (**Figure 5A**). Unstimulated condition was used as negative control to exclude the background response. We utilized the activation induced marker assay (AIM) for quantification of primed CD4^+^ T cells (Dan et al., 2016) during early stages after 2 days of co-cultures. We were able to detect AIM positive (AIM^+^) CD4^+^ T cells co-expressing OX40 and CD25 in all the co-cultures (T+M, T+B, T+B+M) in presence of both antigens, Mtb peptides (**Figure 5B, C**; Unstimulated versus Mtb peptides; T+M, P=0.006; T+B, P=0.01; T+B+M, P=0.0002) and Mtb lysate (**Figure 5B, D**; Unstimulated versus Mtb lysate; T+M, P=0.02; T+B, P=0.03; T+B+M, P=0.003). In response to Mtb peptides stimulation, the magnitude of background subtracted AIM^+^CD4^+^ T cells was more robustly detected in the presence of monocytes supplemented co-cultures, which was significantly higher than those detected in the absence of monocytes in T+B alone (**Figure 5E**; T+M versus T+B, P=0.007; T+B+M versus T+B, P<0.0001). Like Mtb peptides, stimulation with Mtb lysate also showed a similar pattern of robust activation of AIM^+^ CD4^+^ T cells in the T+M and T+B+M co-cultures significantly higher than in the T+B alone (**Figure 5E**; T+M versus T+B, P=0.04; T+B+M versus T+B, P=0.004). We further examined the expression of CD40L molecule on the AIM^+^ CD4^+^ T cells, which was detected in all the co-cultures in response to the stimulation with Mtb antigens (**Figure 5G**). CD40L expressed on activated CD4^+^ T cells function as a co-stimulatory molecule to deliver constant survival and proliferation signals to the APCs like B cells (Elgueta et al., 2009; Garside et al., 1998; Xu et al., 1994). We found that co-cultures containing monocytes (T+M and T+B+M) induced the CD40L expression (CD40L^+^ cells) in AIM^+^ CD4^+^ T cells almost 2-fold higher than the co-culture without monocytes (T+B) in presence of both antigens, Mtb peptides (**Figure 5H**; T+M, 59±6%; T+B, 36±4%; T+B+M, 58±3%) and Mtb lysate (**Figure 5H**; T+M, 65±4%; T+B, 38±5%; T+B+M, 56±4%). The magnitudes of AIM^+^CD4^+^ T cells and CD40L^+^ cells in the co-cultures of T+M and T+B+M also indicated that the presence of B cells in T+M co-culture did not bring significant favor for priming and activation of CD4^+^ T cells at the early stages. Hence, the results suggest that presence of monocytes in T-B co-cultures contributed to an initial priming and activation of antigen-specific T cells during the early stages of co-culture, superior than the B cells. The early efficient priming by monocytes seems to endowed the T cells with strong potential of cognate interactions with B cells and to deliver the vital helper signals in the successive stages of co-culture.

**Figure 5.**
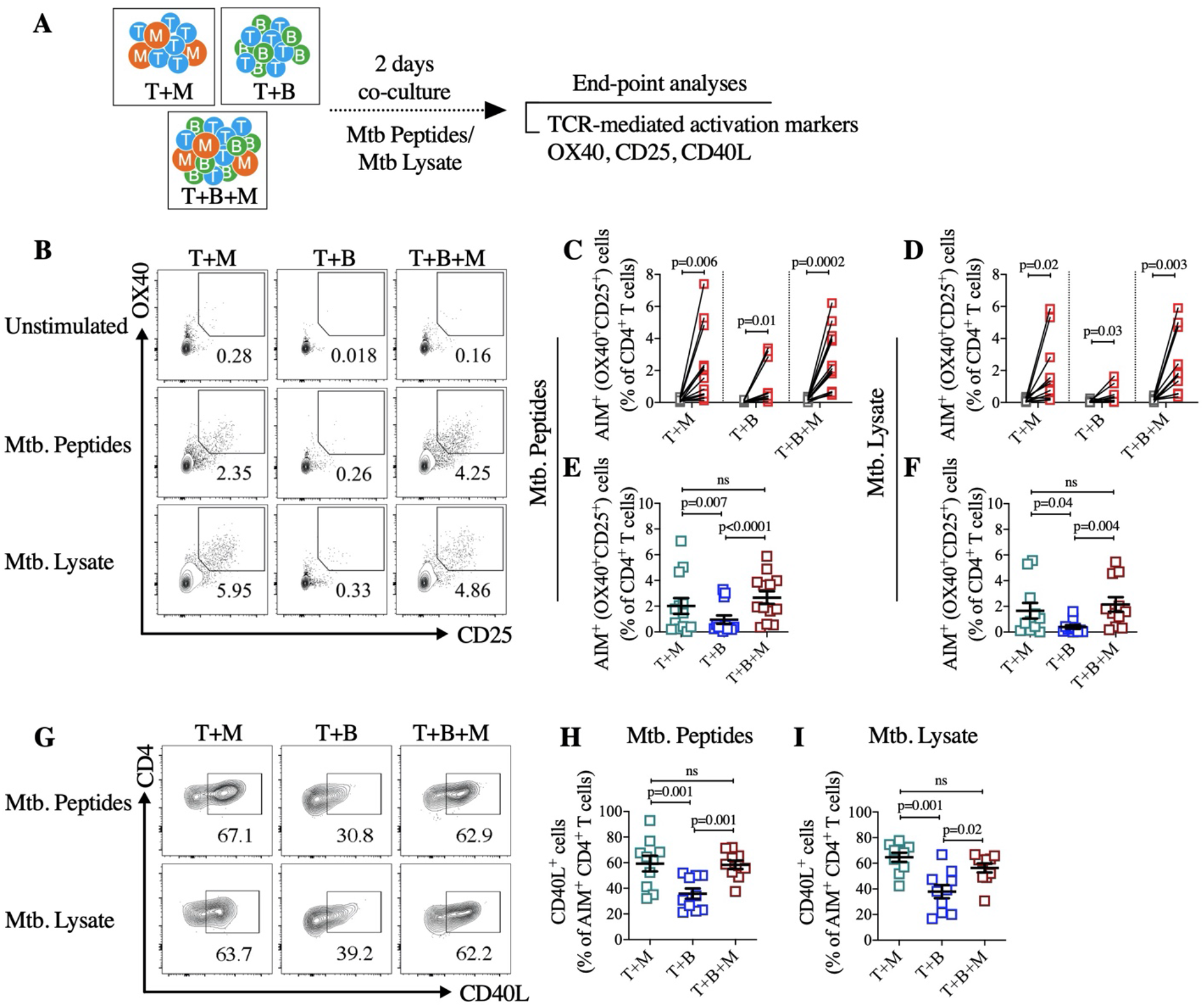
Monocytes act as superior antigen presenting cells for initial priming and activation of antigen-specific memory CD4^+^ T cells during early stage of co-cultures. **(A)** Schematic representation for the analysis of TCR-mediated activation of antigen-specific memory CD4^+^ T cells in presence of monocytes, B cells or combination of both during early stage of co-cultures at Day 2. **(B)** Representative contour plots for the flow cytometric analysis of AIM^+^ (OX40^+^CD25^+^) CD4^+^ T cells (gated in CD4^+^ T cells) in T+M, T+B and T+B+M co-cultures in unstimulated control condition or stimulated conditions with *M. tuberculosis* (Mtb.) peptides and Mtb. whole cell lysate (Mtb. lysate) at Day 2 post co-culture. AIM^+^ CD4^+^ T cells shown in T+M, T+B and T+B+M co-cultures in (**C**) Mtb. peptides and (**D**) Mtb. Lysate stimulated conditions and compared with their unstimulated control conditions. Line connecting grey and red open circles represent the same individual in the unstimulated and stimulated conditions, respectively. Background subtracted AIM^+^ CD4^+^ T cells in (**E**) Mtb. peptides and (**F**) Mtb. lysate stimulated conditions, calculated as AIM^+^ CD4^+^ T cells in stimulated condition minus AIM^+^ CD4^+^ T cells in unstimulated control condition. (**G**) Representative contour plots for the expression of CD40L in AIM^+^ CD4^+^ T cells in T+M, T+B and T+B+M co-cultures in Mtb. peptides and Mtb. lysate stimulations. Percent CD40L cells in AIM^+^ CD4^+^ T cells in T+M, T+B and T+B+M co-cultures in (**H**) Mtb. peptides and (**I**) Mtb. lysate stimulated conditions. Data is represented as mean ± sem, with each dot representing one donor. Data represent the pool of three independent experiments. Statistical comparisons were performed by (C-D) two-tailed paired t test, (E-F and H-I) one-way ANOVA followed by Bonferroni’s multiple comparisons test. ns: non-significant.

### *Monocytes driven enhanced B cell survival is largely mediated by BAFF and* SCGF-β*/CLEC11A*

Survivability of B cells is enhanced in the presence of monocytes. In addition to the efficient early priming of T cells, the enhanced survivability of B cells could be the partial explanation for why monocyte enhanced the T cell-B cell collaboration in presence of specific antigens. Thus, we intended to look into the various factors including growth/survival factors secreted in monocyte supplemented condition. In unstimulated autologous cultures, we measured the chemokines, cytokines and growth factors secreted in the supernatant of B+M co-culture and compared it with the B cell alone condition. We detected significantly higher concentration of chemokines, CXCL1, CCL2, CXCL8 and cytokines, IL-1α, IL-6, TNF-α, IFN-γ and IFN-α2 in B+M co-culture compared with B alone (**Figure 6A, B; Figure S6**). Growth factors like G-CSF, GM-CSF, M-CSF, TRAIL, LIF, MIF and SCGF-β were also detected in higher quantity in presence of monocytes (**Figure 6A, B; Figure S6**). Thus, we investigated the role of survival factors that were detected in substantial concentration in B+M co-culture such as IL-6, SCGF-β and MIF. In addition, we also examined the implication of BAFF that is known to promote survival and proliferation of mature B cells (Batten et al., 2000; Fu et al., 2009; Schweighoffer et al., 2013). We cultured 60,000 CD20^+^ B cells with CD14^+^ monocytes (M) for 6 days in 1:2 ratio (M to B) in unstimulated conditions either in absence of any antibody, isotype antibody or neutralizing antibodies against BAFF, IL-6, SCGF-β and MIF. Blocking the secreted BAFF molecules by neutralizing antibody (αBAFF) significantly reduced the live B cell numbers. Although, individually blocking the secreted IL-6 was not as effective as blocking the BAFF, the SCGF-β blocking using the neutralizing antibody (αSCGF-β) was highly efficient in reducing the B cell survival (**Figure 6D**; count: isotype, 2889±125 versus αBAFF: 1767±92; P<0.0001; αSCGF-β:1776±149; P<0.0001; αIL6: 2140±166; P=0.0001). Besides, blocking of secreted MIF showed a marginal reduction in the count of live B cells (**Figure S7**; isotype, 2645±85 versus αMIF, 2325±124; P=0.01). Thus, the blocking of BAFF and SCGF-β was equally superior in reducing the monocyte dependent enhanced B cell survival, which was higher than the effect of blocking IL-6 on the B cell survival. Interestingly, a significant synergistic effect on the reduction in B cell survival was observed when SCGF-β blocking was combined with the BAFF blocking (**Figure 6D;** count: isotype versus αBAFF+αSCGF-β: 1268±124; P<0.0001). Thus, blocking BAFF was able to reduce the survival of B cells to 56±3%, which was further reduced to 38±5% in the combination blocking in presence of αSCGF-β (Fig. 6E; Isotype vs αBAFF+αSCGF-β: P<0.0001). Combination blocking results suggest that the signaling axis driven by BAFF and SCGF-β is majorly participating in the B cell survival. Altogether, these data revealed that monocytes mediated enhanced B cell survival is majorly supported by BAFF and newly described growth factor SCGF-β/CLEC11A in the monocytes supplemented co-cultures.

**Figure 6.**
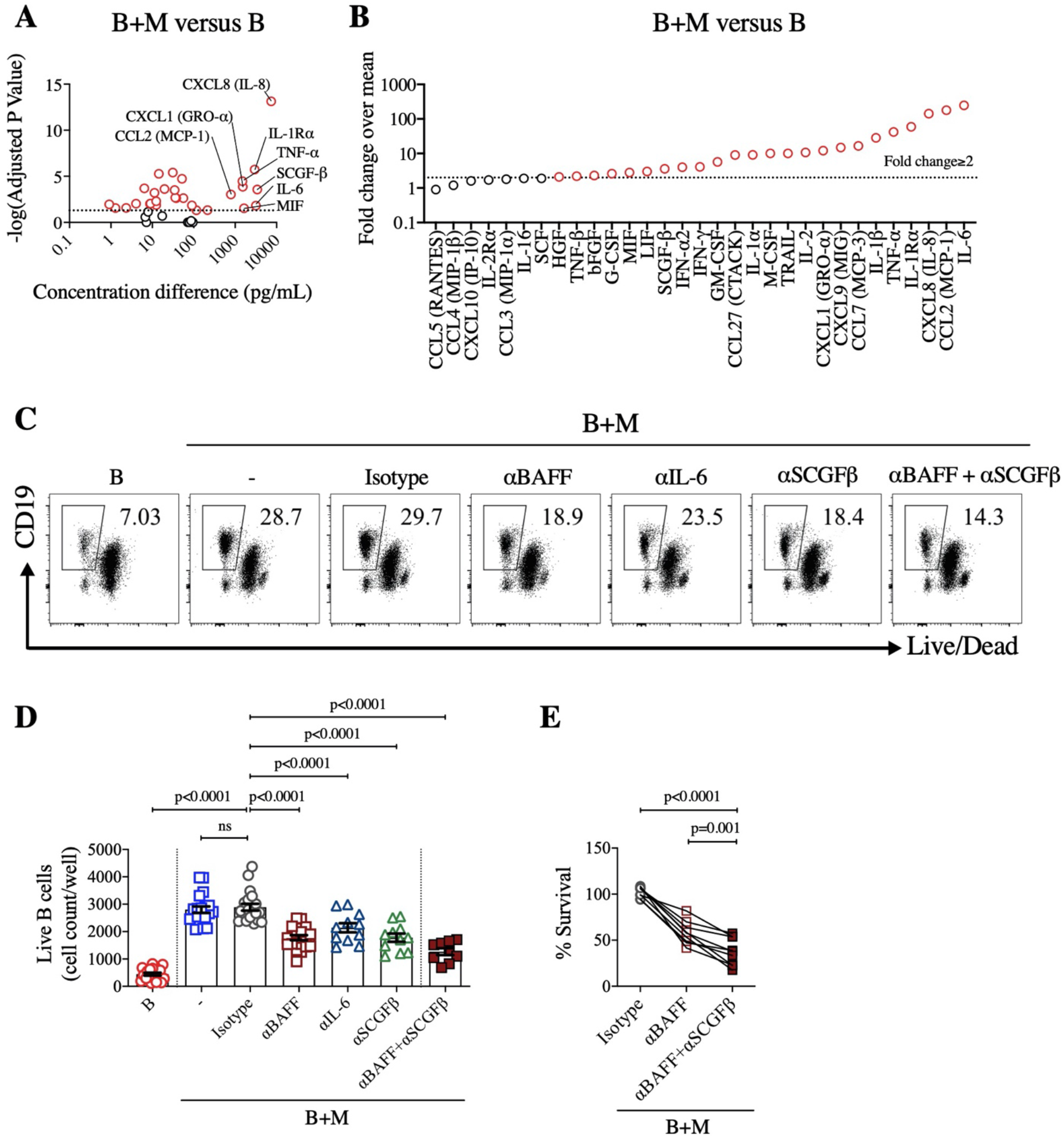
BAFF and SCGFβ are majorly implicated in the monocytes driven enhanced B cell survival. B cells were co-cultured with monocytes in ratio of 1:2 (monocyte to B cells) in unstimulated condition for 9 days and secreted soluble factors in culture-supernatant were quantified using a bead based multiplex immunoassay. (**A**) Shown is the volcano plot representing differentially secreted soluble factors in B+M co-culture versus B alone. The -log10 (adjusted p-value) was calculated by multiple t-test corrected using the Bonferroni-Sidak method plotted against concentration difference (pg/mL) between B+M co-culture and B alone. Dotted horizontal line represent the -log10 (adjusted p-value) corresponding to p=0.05. Red open circles represent the analyte significantly secreted in high quantity in B+M co-culture relative to B alone. (**B**) Shown is the fold-change (calculated by dividing mean level of analyte in B+M with mean level in B alone) of differentially secreted soluble factors. Dotted horizontal line represent the fold change of 2. Red open circles represent the analyte with fold change of ≥2. Data is arepresentative of n=10 donors. For blocking experiments, B cells were co-cultured with monocytes in ratio of 1:2 (monocyte to B cells) in unstimulated condition for 6 days in presence of neutralizing antibodies against BAFF, IL-6, SCGFβ or their combination. For isotype control, mouse IgG was used in the same concentration as neutralizing antibodies. (**C**) Representative dot plots for the flow cytometric analysis of live B cells in B+M co-cultures in presence of isotype (IgG), anti-BAFF, anti-IL-6, anti-SCGFβ, and anti-BAFF+anti-SCGFβ. (**D)** Count of live B cells (live CD19^+^ cells) shown in B alone, B+M co-cultures containing no antibodies (-), isotype (IgG) and neutralizing antibodies (αBAFF, αIL-6, αSCGFβ, and αBAFF+αSCGFβ). (**E)** Percent survival of B cells shown in B+M co-cultures in presence of isotype, αBAFF, and αBAFF+αSCGFβ. Percent survival of B cells was calculated by first normalizing the B cell count with respect to B cell alone (subtracting it from all the wells) and then calculating the percentage over B+M co-culture containing no antibodies (-). Data represent the pool of two independent experiments. Statistical comparisons were performed by (D-E) one-way ANOVA followed by Bonferroni’s multiple comparisons test. ns: non-significant.

## Discussion

The availability of T-cell and B cell co-cultures is instrumental for the functional assessment of CD4^+^ T cells in inducing T-dependent B cell responses. In this study, we have shown that it is possible to examine the quality of CD4^+^ T helper cells in an antigen-specific manner with a greater sensitivity than conventionally used co-culture assays. The most striking finding is the implication of autologous monocytes in providing the opportunity for better collaboration between cognate T cells and B cells. In fact, monocytes provided two most crucial signals in the T-B co-cultures; early priming of CD4^+^ T cells for better T-B cognate interactions, and the survival signals to B cells that was mainly mediated by BAFF and newly identified soluble factor SCGF/CLEC11A.

The robust presence of *M. tuberculosis* (Mtb)-specific memory CD4^+^ T cells in healthy blood donors provided an opportunity to study and optimize the antigen-specific T-B co-cultures. The poor survivability of T and B cells, and the inefficient plasma cell output in Mtb-specific conditions was prominent in the conventional settings of T-B co-cultures. It seems that B cells presenting CD4^+^ peptides do not survive long enough till the Mtb-specific CD4^+^ T cells expand and become armed to deliver the T-dependent helper signals to B cells. Clearly, the conventional settings were lacking essential signals for the survival and efficient cross-talk between T and B cells. The cognate T-B interactions at the T-B border or in the germinal centers are supported by various signals from the myeloid cells and stromal cells (Pereira et al., 2010; Victora and Nussenzweig, 2012). Myeloid cells like monocytes have been implicated in providing survival and expansion support to B cells in various pathologies including the tumor microenvironment. Indeed, supplementation with autologous monocyte supported the B cells in co-cultures to survive more profoundly than with memory CD4^+^ T cells alone. In absence of any exogenous antigen, the B cell survivability was increased by >30 fold in settings of monocyte supplementation. The survival of naïve B cell was prominent than the memory B cells, may be due to the intrinsic capability of memory B cells in higher expression of anti-apoptotic genes (Good et al., 2009; Tangye et al., 2003). The monocyte supplementation effect was clearly visible in *M. tuberculosis* (Mtb)-specific conditions, which provided an opportunity to attain the detectable levels of plasma cell and antibodies in cultures by almost 10-fold higher output than the conventional settings. In contrast to Mtb peptides, Mtb lysate stimulation showed higher expansion of Mtb-specific activated memory CD4^+^ T cells, plasma cells and the secreted antibodies. It could be due to the presence of various antigens in the lysate leading to the larger breadth of expansion in specific B and T cells (Cooper, 2009). Importantly, the monocyte supplementation enhanced the sensitivity and provided consistent detection of plasma cell differentiation and antibody response in the presence of both Mtb peptides and lysate antigens. Monocytes supplementation was also consistently associated with robust expansion of Mtb-specific memory CD4^+^ T cells in presence of both of these Mtb antigens. Importantly, monocytes mediated enhanced B cell response was largely dependent on the productive cognate interaction of CD4^+^ T cells and B cells, as the absence of CD4^+^ T cells or absence of Mtb antigens did not lead to any plasma cell or antibody output from the cultured B cells.

The prominent association of CD4^+^ T cells in robust response to SARS-CoV-2 is widely recognized (Lipsitch et al., 2020; Sette and Crotty, 2021). Many studies including our recent work showed the persistent memory CD4^+^ T cells in COVID-19 recovered donors, mostly targeted towards the SARS-CoV-2 spike protein (Ansari et al., 2021; Braun et al., 2020; Grifoni et al., 2020; Le Bert et al., 2020; Low et al., 2021; Weiskopf et al., 2020). However, the qualitative evaluation of SARS-CoV-2 specific CD4^+^ T cells in providing help to B cells is not attempted. We observed plasma cell differentiation, IgG production and antibody secreting cells (ASCs) in SARS-CoV-2 spike CD4 peptides (CoV-S peptides) stimulated T-B co-cultures. In these T-B co-cultures, we also detected a corresponding expansion of SARS-CoV-2 spike specific CD4^+^ T cells, which can secrete IL-21 to help B cells for their differentiation and functions. These observations supported the presence of spike-specific functional memory CD4^+^ T cells in the COVID-19 recovered donors. However, SARS-CoV-2 spike protein (CoV-S protein) stimulated conventional T-B co-cultures showed no plasma cell differentiation or antibody secretion, which coincided with the failure in generation of ICOS^+^PD1^+^ CD4^+^ T cells. The observed differences in CoV-S peptides and CoV-S protein could be partly explained by the way these antigens are processed and presented to prime the specific CD4^+^ T cells. CoV-S protein needs to be internalized, processed and presented by spike-specific B cells through surface BCRs. In contrast, CoV-S peptides could be directly presented on MHC-II of spike-specific or non-specific B cells leading to vigorous priming of the specific CD4^+^ T cells. However, later condition also leads to prominent bystander B cell expansion as evident with the total IgG and total ASCs responses. Certainly, monocyte supplemented T-B co-cultures overcome these limitations by consistently enhancing the plasma cell output and antibody production in co-cultures stimulated with CoV-S protein and peptides. Moreover, the improved sensitivity in the B-cell output in monocyte supplemented settings was accompanied by the enhanced magnitude of functional activated CD4^+^ T cells, which was associated with enhanced level of TCR directed effector cytokines like IL-2, IFN-γ, IL-4, IL-10, IL-17 and sCD40L (Geginat et al., 2001; Rochman et al., 2009). Monocytes also provided crucial cytokines like IL-15 that seems to promote the survival and proliferation of T cells in these spike-specific co-cultures (Dooms et al., 1998; Geginat et al., 2001). These observations also highlighted the fact that these COVID-19 recovered subjects harbor spike-specific memory CD4^+^ T cells that are capable of inducing B cell differentiation into ASCs and produce IgG during any recall response.

Monocytes and B cells act as antigen presenting cells to prime and activate the memory CD4^+^ T cells (Hong et al., 2018; Hua and Hou, 2020; Randolph et al., 2008; Tacke et al., 2006). However, they differ in their abilities to deliver the secondary activation signals as evident by the constitutive expression of CD86 on all classical monocytes, but not on the B cells. In fact, signals from activated T cells like IL21 is essential for inducing the co-stimulatory ligands on the B cells (Attridge et al., 2014). Besides, monocytes are more effective antigen presenting cells than B cells for activating all sub-types of human CD4^+^ T cells (Beck et al., 1996). Monocytes majorly secrete IL-1β and IL-6, which could also act as secondary activation signals for T cells (Ben-Sasson et al., 2009). Indeed, we demonstrate that monocytes are superior to B cells in priming and activating the Mtb-specific CD4^+^ T cells in response to Mtb peptides or lysate antigens, during early stages of co-culture. These activated CD4^+^ T cells expressed higher level of CD40L molecule, which is a key ligand for stimulating the B cells via CD40 axis (Elgueta et al., 2009). Intrinsic potential of T cells to help plasma cell differentiation is secondary to the initial priming via TCR and co-stimulatory molecules and therefore, only activated T cells can deliver their effector functions to B cells. Thus, pre-activation of antigen specific CD4^+^ T cells is necessary in T-B co-cultures to establish the cognate adhesion and activation of the B cells. It was clearly evident in the absence of monocytes supplementation, where T-B co-cultures showed poor activation of Mtb specific T cells at both early and later stages of co-cultures. Although, B cells do not provide efficient signals to prime and activate CD4^+^ T cells in early co-culture period, they could receive T-dependent helper signals from the optimally activated CD4^+^ T cells expressing co-stimulatory molecules in the successive stages of co-cultures.

Furthermore, the enhanced survivability of B cells in presence of monocytes helped them to remain available for interaction and establishing the cognate ligation with armed T cells. Survivability of B cells is an important aspect of T-B co-cultures in order to retain significant number of antigen-specific B cells for the cognate T-cell engagement. In *ex vivo* settings, the specific requirement to keep survive B cells without any antigen is not fully understood. However, B cells survive the *ex vivo* cultures in the presence of stimulation with synthetic ligands, such as TLR7/8 and TLR9 agonists, which help B cells to activate and differentiate into plasma cells in antigen non-specific manner (den Hartog et al., 2018; Jahnmatz et al., 2013). Also, R848 and CpG augment the non-specific proliferation of the B cells in presence of T cell derived factors, IL-21 and CD40L (Auladell et al., 2019; Franke et al., 2020). Besides, enhanced survival and proliferation of B cells was associated with the accumulated monocytes in the tumor microenvironment (Epron et al., 2012). Our study clearly demonstrate that monocyte supplementation provides crucial signals for the enhanced survivability of B cells in the ex vivo co-cultures, which is mainly mediated by the growth factors BAFF and SCGF/CLEC11A. Although, IL6 has been shown to induce the growth-responsive genes in activated B cells (Tosato et al., 1988), it was not superior than BAFF and SCGF-β in enhancing the survivability of B cells in monocyte supplemented co-cultures. Similarly, MIF that seems to impact the B cell survival via CD74/CD44 signaling axis (Gore et al., 2008) has not shown any prominent effect on the B cell survival in monocyte-supplemented cultures. In fact, BAFF seems to play important role in B-cell survival in the monocyte supplemented co-cultures. BAFF is mainly produced by the myeloid cells like monocytes and dendritic cells (Craxton et al., 2003; Nardelli et al., 2001). The implication of BAFF/BAFF-R in survival and maturation of peripheral B cells is well established (Muller-Winkler et al., 2021; Rauch et al., 2009). It’s possible that in monocyte-supplemented conditions, the monocytes provided this important factor to the cultured B cells and promoted their survival via the NF-kB2 signaling axis and via increasing the anti-apoptotic response (Clybouw et al., 2011; Craxton et al., 2005). Moreover, early priming of T cells by monocytes further enhanced the availability of BAFF in co-cultures via boosting monocytes through effector cytokines like IL-10 and IFN-γ(Mueller et al., 2007).

The striking finding of our study is the identification of newly described growth factor SCGF/CLEC11A, as one of the crucial factors for supporting survival of blood B cells. The SCGF was as efficient as BAFF in providing survival signals to the B cells and showed substantial synergy with BAFF in effectively regulating the B cells survival. SCGF is a sulfated glycoprotein that is expressed abundantly in skeletal tissues including bone marrow and also at a low level in lymphoid tissues like spleen (Bannwarth et al., 1998; Hiraoka et al., 2001; Perrin et al., 2001). SCGF was identified as a growth factor that promotes proliferation and survival of hematopoietic stem/progenitor cells. However, SCGF has never been investigated in the context of biology and function of lymphoid cells. A very high concentration of SCGF-β in monocyte supplemented cultures clearly suggest that human monocytes are also capable of secreting this growth factor. The α11β1 integrin is reported as the putative receptor for SCGF on the stromal cells (Shen et al., 2019). Whether it’s the same receptor on B cells or other receptors are implicated in SCGF mediated signaling needs to be determined. In our monocyte supplemented cultures, it’s plausible that SCGF-β is promoting the B-cell survival by inducing activation of extracellular signal– regulated kinase (ERK) pathway like BAFF (Craxton et al., 2005) or via Wnt-mediated activation signaling through its putative integrin receptors (Shen et al., 2019). However, other mechanisms may also contribute to the SCGF mediated survival in the B cells. Thus, in-depth studies are warranted to establish the role of this newly described growth factor in B cell survival and function.

In summary, our study described a highly efficient method for qualitative evaluation of memory CD4^+^ T cells in an antigen-specific manner. We demonstrate that monocytes contributed in enhancing the propensity of cognate T-B interactions by providing early activation signals to the T cells, and via improving the survivability of B cells. Our finding on the implication of SCGF/CLEC11A in B cell survival highlights the crucial role for this growth factor in B cell development and function. Although effective COVID-19 vaccines have been rolled out, the trials for more broadly protective vaccines will continue. This is an unprecedented situation that demands for multiple immunologic parameters for the evaluation of vaccines in early phase of trials as well as after the roll out. Because the induction of humoral response depends on the CD4^+^ T cells, it’s important to utilize B cell help function of CD4^+^ T cells as a correlate of protective adaptive immunity. The higher sensitivity and the ease of using various type of antigen overcome the existing limitations in widespread use of antigen-specific T-B co-culture assays. Thus, the hereby described method provides an opportunity for the assessment of vaccine induced functional memory responses as well as for studying the T-B cross talk and productive outcome in infection and vaccination.

## Methods and Details

### Human Blood Sample

The study was approved by the Institutional Review Boards of National Institute of Immunology (NII) and All India Institute of Medical Sciences (AIIMS), New Delhi, India. Informed consent was obtained from all the donors/subjects during the enrolment. For *Mycobacterium tuberculosis* (Mtb) specific assays, peripheral blood buffy coats from healthy adult donors were collected from blood bank at the AIIMS, New Delhi, India. For SARS-CoV-2 specific assays, peripheral blood from COVID-19 recovered adult subjects were collected at AIIMS, New Delhi, India. The characteristics of participants is provided in the Supplementary Table 1.

### PBMCs Isolation and Cell Sorting

All peripheral blood samples were collected in CPD blood bags for healthy blood donors and K3 EDTA vacutainers for COVID-19 recovered subjects. Peripheral blood mononuclear cells (PBMCs) were isolated by Ficoll-Paque PLUS (GE Healthcare Life Sciences) density gradient medium and cryopreserved in multiple aliquots in FBS (Gibco) containing 10% DMSO (Thermo-Fisher). PBMCs were revived in pre-warmed HyClone RPMI 1640 (Cytiva Life Sciences) and treated with Dnase I (StemCell) at 50μg/mL for 15 minutes at 37ºC. Revival viability of PBMCs was ≥90% as measured by PI/AO staining in the LUNA-FL automatic cell counter (Logos Biosystems Inc., USA). For all the autologous assays, PBMCs after revival were surface stained with antibody cocktail for 15-20 minutes at 4ºC in dark: fixable viability dye efluor 506, anti-CD4 AF700 (RPA-T4), anti-CD45RO FITC (UCHL1), anti-CD14 PE (MΦP9), anti-CD20 PE-Cy7 (2H7). Memory CD4^+^ T cells were sorted from PBMCs as live CD14^-^CD20^-^CD4^+^CD45RO^+^ cells. CD14^+^ monocytes and CD20^+^ B cells were sorted from PBMCs as live CD14^+^CD20^-^ and CD20^+^CD14^-^ cells, respectively. All the cells, T cells, B cells and monocytes were sorted in RPMI 1640 (Gibco) medium containing 50% FBS on a BD FACSAria Fusion flow cytometer (BD Biosciences) at intermediate flow rate, using a 70μm nozzle. The full FACS sorting scheme is shown in Figure S1. Samples with the purity of all the cell type ≥95% were included for the assays.

### Monocyte Supplemented CD4^+^ T cell and B cell (T-B) Co-cultures

FACS sorted memory CD4^+^ T cells were co-cultured with autologous CD20^+^ B cells (60,000 cells each per well) in 1:1 ratio (T to B) in absence (T+B) or presence of autologous CD14^+^ monocytes (T+B+M) (30,000 cells per well) for 9 days or as mentioned in the respective figure legend. For T cell + Monocytes (T+M) or B cell + Monocytes (B+M) co-cultures, memory CD4^+^ T cells or B cells (60,000 cells each per well) were co-cultured with autologous monocytes (30,000 cells per well). Cells were seeded in U-bottom 96-well plates (Nunc, Thermo) in 200μL per well in AIM-V serum-free medium (Gibco) with no exogenous stimulation (Unstimulated control) or stimulation with *M. tuberculosis* specific pool of CD4^+^ T cell peptides (Mtb peptides, 1μg/mL) or *M. tuberculosis* strain H37Rv whole cell lysate (Mtb lysate, 10μg/mL) (BEI Resources). For T-B co-cultures in COVID-19 recovered subjects, cells were kept unstimulated or stimulated with SARS-CoV-2 spike specific pool of CD4^+^ T cell peptides (CoV-S peptides, 1μg/mL) or SARS-CoV-2 full length Spike protein (CoV-S protein, 10μg/mL) (Native Antigen, UK). After 9 days of co-culture, phenotyping and cell count of activated CD4^+^ T cells and plasma cells were determined by flow cytometry. Simultaneously, co-culture supernatants were stored at -80ºC for the measurement of secreted IgG concentration by ELISA or cytokine concentration by bead-based multiplex ELISA. In some experiments, after 9 days of co-culture, IgM and IgG ASCs or IL-21 SFCs were measured by FluoroSpot and ELISpot, respectively.

### B cell/T cell Survival and Blocking Assays

FACS sorted CD20^+^ B cells or memory CD4^+^ T cells (60,000 cells each per well) were cultured either alone or co-cultured with autologous CD14^+^ monocytes (30,000 cells per well). Cells were seeded in U-bottom 96-well plates (Nunc, Thermo) in 200μL per well in AIM-V serum-free medium (Gibco) with no exogenous stimulation (unstimulated) for 9 days. After 9 days of incubation, cells were analyzed for the count of live B cells and live T cells by flow cytometry. At the same time, cell culture supernatants were stored at -80ºC for the measurement of cytokine concentration by bead-based multiplex ELISA. For the titration of monocyte to B cell ratio, B cells (60,000 cells per well) were co-cultured with autologous monocytes in various M to B ratios: 1:4, 1:2 and 1:1 in unstimulated condition for 9 days. For the blocking experiments, sorted CD20^+^ B cells (60,000 cells per well) were cultured either alone (B alone) or with autologous CD14^+^ monocytes (B+M) (30,000 cells per well) in unstimulated condition for 6 days. Soluble growth factors/cytokines in B+M co-cultures were blocked using 10μg/mL of following neutralizing antibodies: purified anti-BAFF (#AF124) and anti-SCGF-β (#MAB1904) (R&D Systems), purified anti-IL-6 (MQ2-13A5) and anti-MIF (10C3) (BioLegend). For isotype control, 10μg/mL purified mouse antibody IgG (4G2) was used in B+M co-cultures. After 6 days of incubation, cells were analyzed for the count of live B cells by flow cytometry.

### Flow Cytometric Analyses

Phenotypic analysis and cell count measurement of T cells and B cells at different time points of T-B co-cultures were performed by flow cytometry. Briefly, cells were surface stained with antibody cocktail in U-bottom 96-well plates (Nunc, Thermo) in FACS buffer (PBS containing 2% FBS) for 30-40 minutes at 4ºC in dark. Antibody cocktail contained the following antibodies: fixable viability dye efluor 506, anti-CD3 APC-Cy7 (HIT3a), anti-CD4 AF700 (RPA-T4), anti-CXCR5 AF647 (RF8B2), anti-ICOS FITC (C398.4A), anti-PD-1 PE (EH12.2H7), anti-CD19 BV786 (HIB19), anti-CD27 PE-Dazzle594 (M-T271), anti-CD20 PE-Cy7 (2H7), anti-CD38 PE-Cy5 (HIT2). Following the surface staining, cells were washed with FACS buffer and resuspended in 200μL FACS buffer. The full gating scheme for analysis of plasma cells and activated CD4^+^ T cells is shown in Figure S1. All the samples were acquired on BD LSR Fortessa flow cytometer (BD Biosciences). Data were analyzed using FlowJo 10.3.0. Phenotyping of monocytes and B cells were performed immediately after reviving PBMCs from healthy donors. PBMCs were surface stained in FACS buffer with following antibody cocktail: fixable viability dye efluor 506, anti-CD4 AF700 (RPA-T4), anti-CD14 BV786 (M5E2), anti-CD20 APC-efluor780 (2H7), anti-CD27 PE-Dazzle594 (MT-271), anti-IgD FITC (IA6-2), anti-IgM APC (MHM-88), anti-HLADR BV650 (L243), anti-CD80 PE (L307.4), anti-CD86 PE-Cy7 (2331(FUN-1)). Gating strategy for phenotyping of monocytes and B cell subsets is shown in Figures S5.

### Activation Induced Marker (AIM) Assay

Early stage priming and TCR-downstream activation of antigen-specific memory CD4^+^ T cells was measured by AIM assay. In this assay, activated CD4^+^ T cells were detected by phenotyping concurrent expression of OX40 and CD25. Healthy donor PBMCs were revived and surface stained for FACS sorting of memory CD4^+^ T cells, CD20^+^ B cells and CD14^+^ monocytes as mentioned previously, with the following antibody cocktail: fixable viability dye efluor 506, anti-CD4 AF700 (RPA-T4), anti-CD45RO FITC (UCHL1), anti-CD14 BV786 (M5E2), anti-CD20 APC-efluor780 (2H7). Sorted memory CD4^+^ T cells (60,000 cells per well) were co-cultured with either CD14^+^ monocytes (30,000 cells per well) or CD20^+^ B cells (60,000 cells per well) or both monocytes and B cells in autologous manner. Cells were seeded in U-bottom 96-well plates (Nunc, Thermo) in 200μL per well in AIM-V serum-free medium for 2 days. Cells were kept either unstimulated (no exogenous stimulation) or stimulated with Mtb peptides (1μg/mL) or Mtb lysate (10μg/mL) for stimulation of antigen specific CD4^+^ T cells. After incubation, cells were stained with the following antibody cocktails: fixable viability dye efluor 506, anti-CD3 BV650 (UCHT1), anti-CD4 AF700 (RPA-T4), anti-OX40 PE-Cy7 (Ber-ACT35), anti-CD25 FITC (M-A251) and anti-CD20 PE-Cy7 (2H7). Following the surface staining, cells were washed with FACS buffer, fixed and permeabilized with Foxp3/Transcription factor staining buffer set (bioscience) followed by intracellular staining with anti-CD40L APC (). After staining cells were washed and re-suspended in 200μL FACS buffer. Samples were acquired on BD LSR Fortessa flow cytometer. Data were analyzed using FlowJo 10.3.0.

### Enzyme-Linked Immunosorbent Assay (ELISA)

We used in-house developed indirect ELISA for measurement of SARS-CoV-2 spike specific IgG antibody in the co-culture supernatants as described previously (Ansari et al., 2021). Maxisorp ELISA 96-well plates (Nunc, Thermo) were coated with 50μl/well SARS-CoV-2 full length Spike protein (CoV-S protein, 1μg/mL) (Native Antigen, UK) in PBS for overnight at 4ºC. After wash, the plates were blocked with blocking buffer (PBS containing 3% Skim milk, 0.05% Tween-20) and incubated at room temperature (RT) for 2 hours. After 5x washing, 50μl/well neat cleared supernatants were added in duplicates and incubated at RT for 1.5 hours. After 5x washing, Goat anti-human IgG conjugated with HRP (Southern Biotech) was added and plates were incubation at RT for 1 hour. For calculating the concentration of SARS-CoV-2 spike specific IgG antibody, purified SARS-CoV-2 neutralizing antibody (CR3022) was used for plotting the standard. For measurement of the total IgG, we used another in-house developed sandwitch ELISA. Plates were coated with 100μl/well purified Goat anti-human Ig Fab unlabeled (0.5μg/mL) (Southern Biotech). Samples were appropriately diluted in diluent buffer (PBS containing 1% Skim milk, 0.05% Tween-20) and the concentration of human IgG was calculated by plotting standard with the purified human IgG. Goat anti-human IgG Fc-HRP (Southern Biotech) was used to detect the bound human IgG. The reaction was developed by adding 100μl/well OPD peroxidase substrate (Sigma) for 10 minutes in dark at RT. The reaction was stopped by adding 50μl/well of 2N hydrochloric acid (HCl), followed by optical density (OD) measurement at 492 nm using MultiskanGO ELISA reader (Thermo-Fisher).

### Bead-Based Multiplex Cytokine Immunoassay

Bio-Plex Pro Human Cytokine Panel 14-Plex and Pro Human Th17 3-plex set (BIO-RAD) was used to quantitate the concentration of soluble cytokines in supernatants from 9 days co-cultures, T+B and T+B+M wells stimulated with CoV-S protein. Bio-Plex Pro Human Cytokine Screening 48-Plex Panel was used to measure the quantity of secreted factors in supernatants from 9 days co-cultures, B+M and B alone wells kept unstimulated. The cell free supernatants were stored at -80ºC until thawed for the quantifications of secreted factors. The multiplex cytokine immunoassays were performed as per the manufacturer’s instructions.

### FluoroSpot/ELIspot Assay

SARS CoV-2 spike specific antibody secreting cells (ASCs) were detected by dual-fluorochrome FluoroSpot assay. Multiscreen IPFL FluoroSpot plate (Mabtech) was charged with 35% ethanol and coating was done with SARS-CoV-2 full length Spike protein (CoV-S protein, 5μg/mL) (Native Antigen, UK) for overnight at 4ºC. For detection of total ASCs, plates were coated with anti-human capture mAbs MT91/145 (for IgG, 15μg/mL) and MT11/12 (for IgM, 15μg/mL) (Mabtech). Plates were washed and blocked with AIM-V medium containing 10% FBS for at least 30 minutes at RT. After 9 days of co-culture, cells from CoV-S peptides stimulated wells were seeded at 4/5^th^ dilution for SARS-CoV-2 spike specific ASCs and 1/5^th^ dilution for total ASCs. Cells were incubated at 37ºC for 16 hours. Cells were discarded and plates were washed 5 times with PBS. For detection of ASC spots, anti-human IgG-550 (MT78/145) and IgM-640 (MT22) detection antibodies (Mabtech) were added and incubated for 2 hours at RT in dark. Plates were washed 5 times with PBS Fluorescence enhancer-II (Mabtech) and incubated for 15 minutes at RT. ASC spots were read on AID *vSpot* Spectrum Elispot/Fluorospot reader system using AID Elispot software version 7.x. ASC counts were calculated by multiplying the detected counts with dilution factor. IL-21 secreting cells were detected by human IL-21 ELISpot (3540; Mabtech). ELISpot plate (MSIPS4510; Millipore) was charged with 35% ethanol and coated with 10μg/mL of coating mAb MT216G (Mabtech), followed by overnight incubation at 4ºC. Plates were washed and blocked as mentioned above. After 9 days of co-culture, cells from CoV-S peptides stimulated wells were seeded either unstimulated or stimulated with CoV-S peptides (1μg/mL). Cells were incubated for 24 hours at 37ºC. Plates were washed 5 times with PBS and incubated with 0.25μg/mL detection mAb MT21.3m-biotin (Mabtech) for 2 hours at RT. After incubation, plates were developed as per the manufacturer’s instructions. IL-21 spot forming cells (SFCs) were read as stated above. SFC counts were calculated by multiplying the detected counts with dilution factor.

### Data Visualization and Statistics

In all the experiments, data are represented as the mean±s.e.m. The significance of the differences between the groups was analyzed with two-tailed paired t-test, two-tailed Mann-Whitney test, two tailed Wilcoxon matched-pairs signed rank test, one-way ANOVA followed by Bonferroni’s multiple comparisons test, multiple t tests corrected using specified methods. P values < 0.05 were considered statistically significant. Statistical analyses and data visualization were performed with the GraphPad Prism software version v8.4.0.

## Data Availability

All data are available in the main text or the supplementary materials.

## Acknowledgments

We are thankful to all the patients and blood donors for generous support in this study. The Mycobacterium tuberculosis, Strain H37Rv, Whole Cell Lysate (NR-14822A) was obtained through BEI Resources, NIAID, NIH. This work was financially supported by the Science and Engineering Research Board, DST grant (IPA/2020/000077) to NG, Department of Biotechnology grant (BT/PR30223/MED/2018) to NG and the core grant of National Institute of Immunology. Further support provided from NIH contract 75N9301900065 (to AS, DW) and NIH grant U01 (U01AI141995-03) to AS.

## Author contributions

Conceptualization and supervision, NG; Formal Analysis, AA, NG; Investigation, AA, SS; Enrolment and sample collection, BPJ, AsS and PC; Resources, AS, DW; Writing, AA, NG; Funding acquisition, NG, AS.

