## Supplementary Data for "A highly efficient T-cell immunoassay provides assessment of B cell help function of SARS-CoV-2 specific memory CD4^+^ T cells"

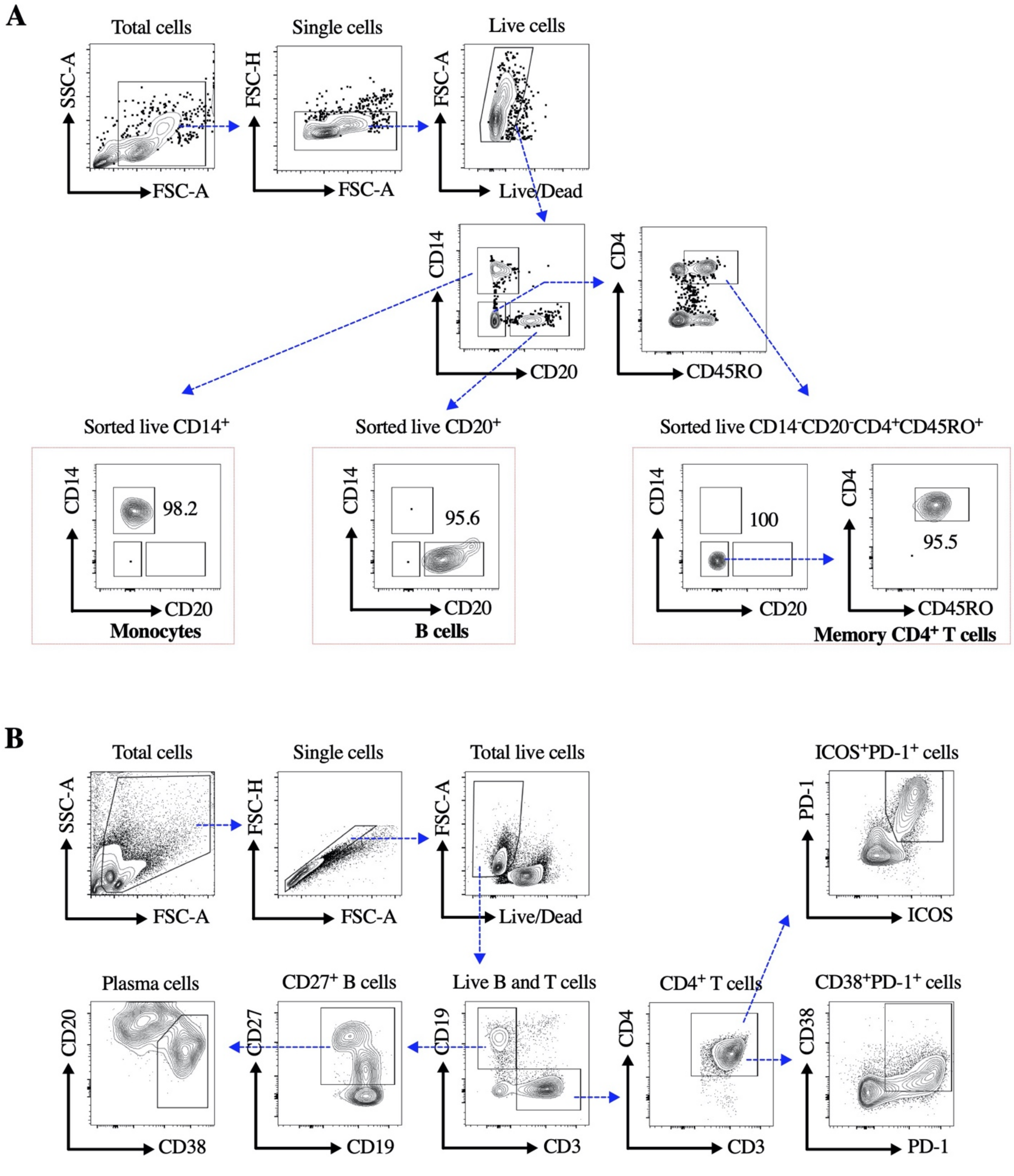

**Supplementary Figure S1. Gating schemes for FACS sorting and flow cytometric analysis in T-B co-cultures.**

(**A**) Representative contour plots for the FACS sorting of monocytes (live CD14$^{+}$CD20$^{-}$ cells), B cells (live CD20$^{+}$CD14$^{-}$ cells) and memory CD4$^{+}$ T cells (live CD14$^{-}$CD20$^{-}$CD4$^{+}$CD45RO$^{+}$cells). (**B**) Representative contour plots for the flow cytometric analysis of plasma cells and activated CD4$^{+}$ T cells.

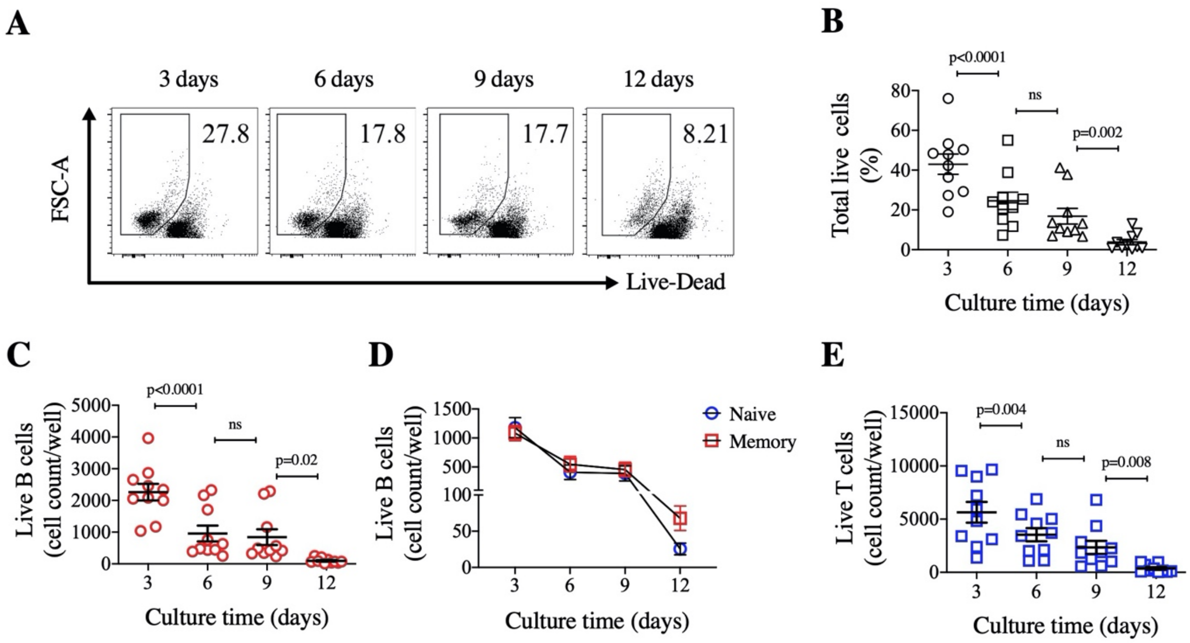

**Supplementary Figure S2. Time kinetics of survivability of B and T cells in T-B co-cultures.**
(**A**) Representative dot plots for the flow cytometric analysis of total live cells in T-B co-culture on various days up to 12 days in absence of any exogenous antigen (unstimulated). (**B**) Percent population of total live cells on various days up to 12 days in unstimulated conditions. (**C**) Shown is the count of live B cells ($CD19^{+}CD3^{-}$ cells gated in live cells) in T+B co-culture on various days in unstimulated condition, and (**D**) Live B cell count of naïve ($CD27^{-}$ B cells) and memory ($CD27^{+}$ B cells) compartments shown in T+B co-culture on various days in unstimulated condition. (**E**) Count of live $CD4^{+}$ T cells ($CD4^{+}CD3^{+}$ cells gated in live cells) in T-B co-culture on various days in unstimulated condition. Data is represented as mean ± sem, with each dot representing one donor. Data represent the pool of two independent experiments. Statistical comparisons were performed by (B, C and E) one-way ANOVA followed by Bonferroni's multiple comparisons test. ns: non-significant.

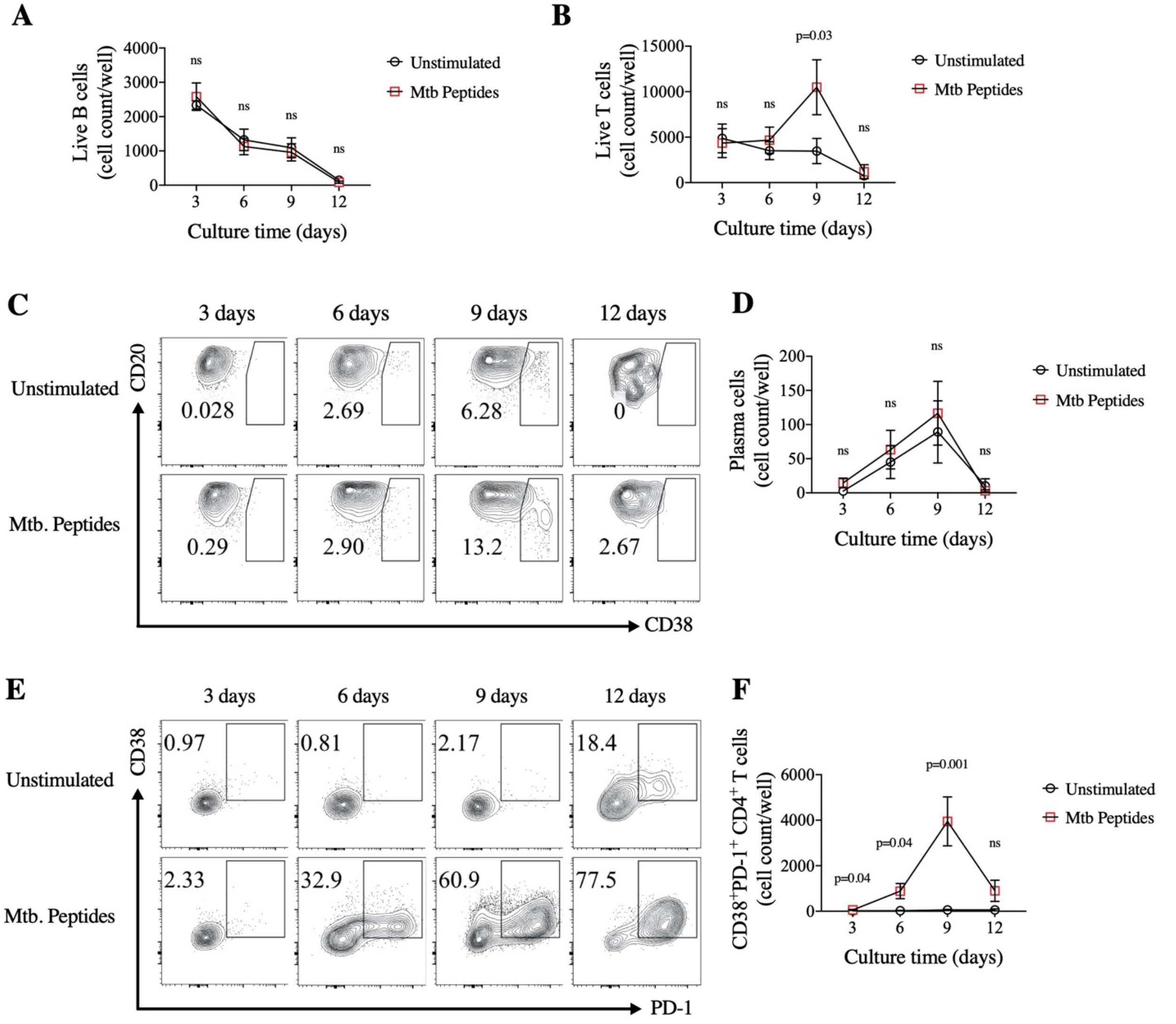

**Supplementary Figure S3. Time kinetics of plasma cells differentiation and activation of T cells in antigen-specific T-B co-cultures.**

(**A**) Live B cell count and (**B**) live $CD4^+$ T cell count shown in T-B co-culture on various days up to 12 days in *Mycobacterium tuberculosis* (Mtb.) peptides stimulated condition (red circle) and compared with their unstimulated control condition (grey circle). (**C**) Representative contour plots for the flow cytometric analysis of plasma cells ($CD20^{lo}CD38^{hi}$cells gated in $CD27^+$ B cells) in T-B co-cultures on various days up to 12 days in unstimulated control condition and Mtb. peptides stimulated condition. (**D**) Count of plasma cells shown in T-B co-culture on various days in unstimulated control condition (grey circle) or in Mtb. peptides stimulated condition (red circle). (**E**) Representative contour plots for the flow cytometric analysis of $CD38^+PD\text{-}1^+CD4^+$ T cells (gated in $CD4^+$T cells) in T-B co-culture on various days in unstimulated control condition and Mtb. peptides stimulated condition. (**F**) Count of $CD38^+PD\text{-}1^+CD4^+$ T cells shown in T-B co-culture on various days in unstimulated control condition (grey circle) or in Mtb peptides stimulated condition (red square). Data is represented as mean ± sem of the results from total 8 donors in two independent experiments. Statistical comparisons were performed by two-tailed paired t test. ns: non-significant.

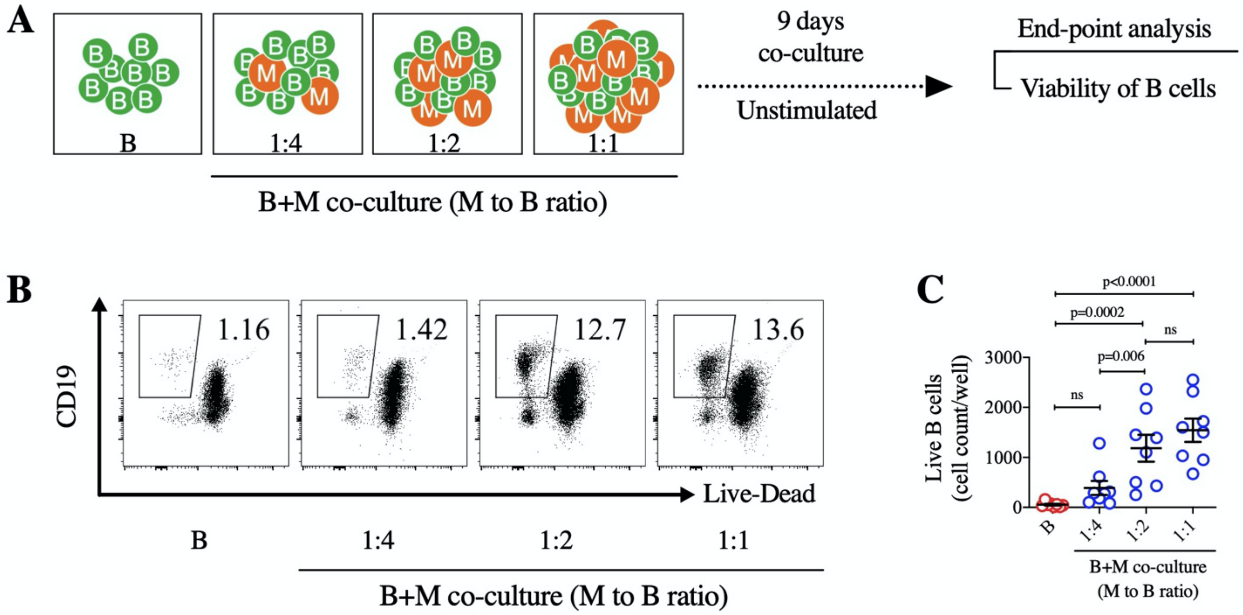

**Supplementary Figure S4. Titration of Monocyte supplementation for B cell survival.**
(**A**) Schematic representation for the titration of monocytes to B cell ratio. (**B**) Representative dot plots for the viable B cells (live $CD19^{+}$ cells) and (**C**) Live B cell count was shown in B alone and co-culture of B cells with monocytes in various M to B ratio of 1:4, 1:2 and 1:1 in unstimulated condition after 9 days of culture. Data is represented as mean ± sem of the results from 8 donors. Statistical analysis was performed by one-way ANOVA and corrected using Bonferroni's multiple comparisons test. ns: non-significant.

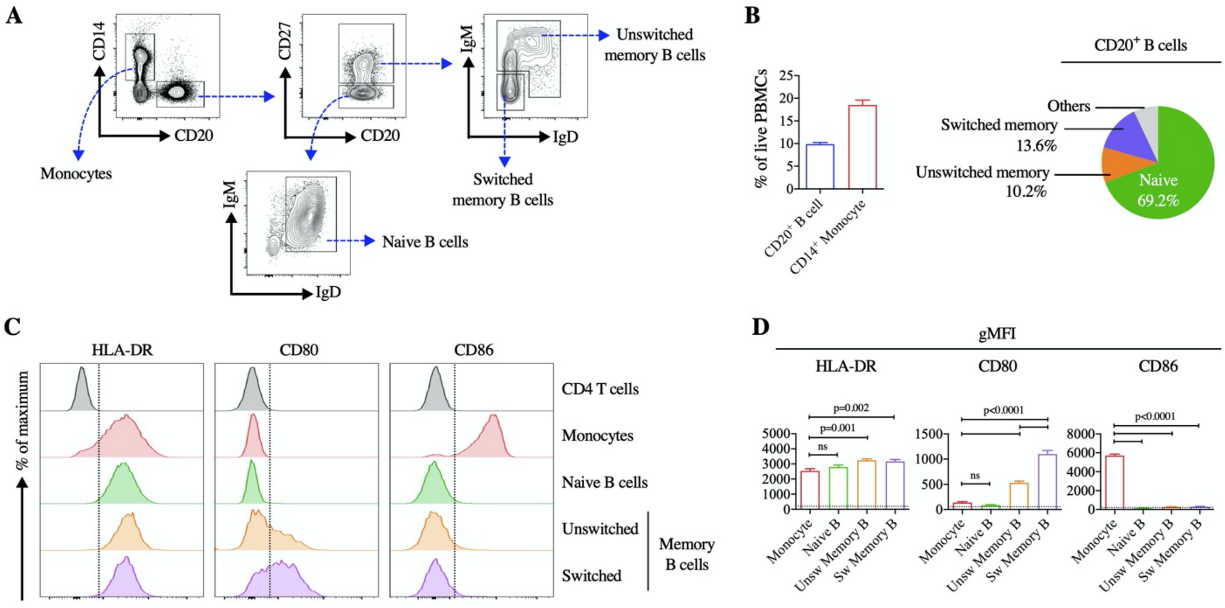

**Supplementary Figure S5. Expression analysis of molecules involved in antigen presentation and T cell activation on B cells and Monocytes, prior to co-culture.**

(**A**) Gating strategy for identifying CD14+ monocytes and various subsets in CD20+ B cells from PBMCs. (**B**) percent of CD20+ B cells and CD14+ monocytes were shown in live PBMCs (*left*). Pie chart showing (mean) proportion of various subsets in CD20+ B cells. (**C**) Representative histogram plots for expression of HLA-DR, CD80 and CD86 on CD4 T cells, monocytes, naïve B cells, unswitched and switched memory B cells. (**D**) Geometric mean fluorescence intensity (gMFI) of HLA-DR, CD80 and CD86 on monocytes, naïve B cells, unswitched and switched memory B cells from fresh PBMCs. Data is represented as mean ± sem of the results from n=8 donors. Statistical analysis was performed by one-way ANOVA and corrected using Bonferroni's multiple comparisons test. ns: non-significant.

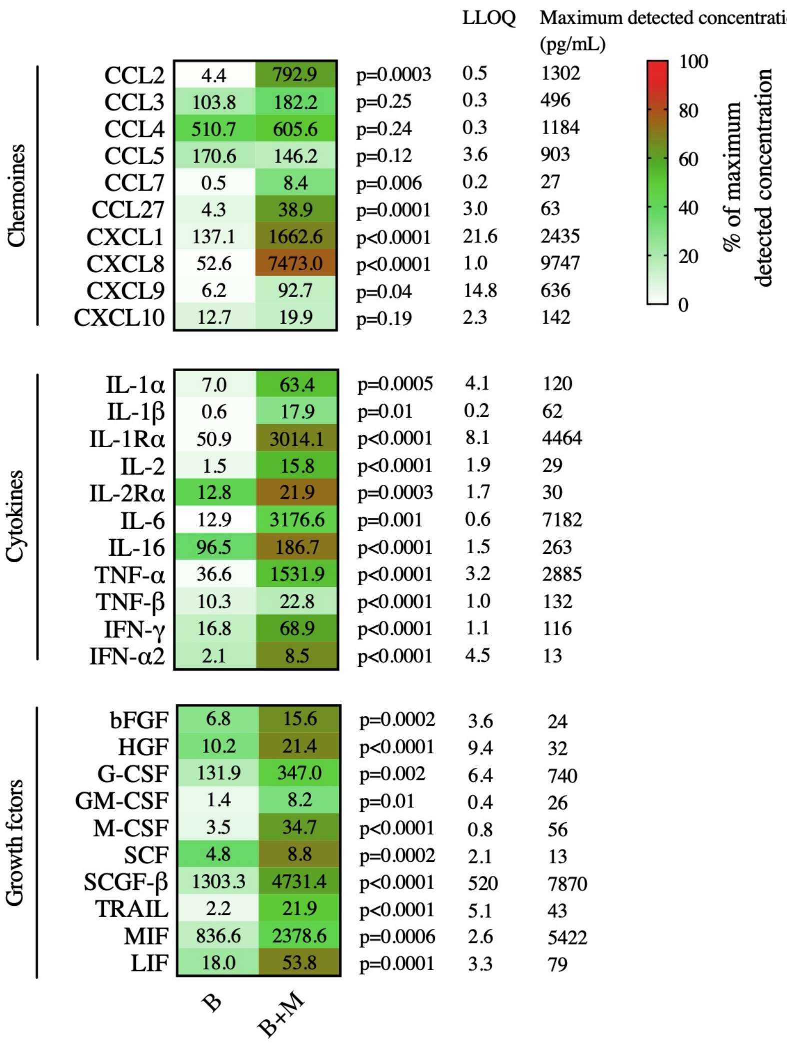

**Supplementary Figure S6. Secreted soluble factors in co-culture of B cells with monocytes.**
B cells were co-cultured with monocytes in ratio of 1:2 (monocyte to B cells) in unstimulated condition for 9 days and secreted soluble factors in culture-supernatant were quantified using a bead-based multiplex immunoassay. Shown is the heat map of the levels of secreted soluble factors in B alone or B+M co-culture. Values in the heat map represent the mean concentrations of the analyte. Heat color scale represents the mean percentage of the maximum detected concentration. Maximum detected concentration of each analyte detected in any of the group was considered as 100%. LLOQ refers to as the lowest limit of quantification. Data is a representative of n=10 donors. Statistical comparisons were performed by two-tailed paired t test.

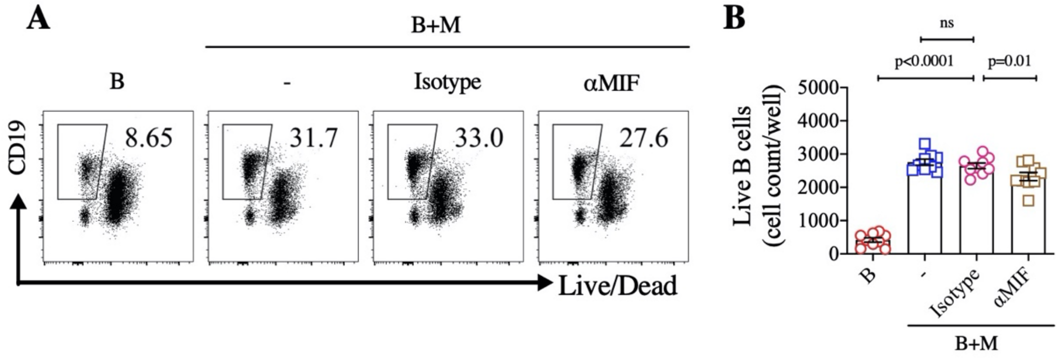

**Supplementary Figure S7. Blocking of MIF in B+M co-culture.**

B cells were co-cultured with monocytes in ratio of 1:2 (monocyte to B cells) in unstimulated condition for 6 days in presence of neutralizing antibodies against MIF. For isotype control, mouse IgG was used in the same concentration as neutralizing antibody. (**A**) Representative dot plots for the flow cytometric analysis of live B cells (live $CD19^+$ cells) in B alone, B+M co-cultures containing no antibodies (-), isotype (IgG), anti-MIF (αMIF). (**B)** Count of live B cells shown in B alone, B+M co-cultures containing no antibodies (-), isotype (IgG) and neutralizing antibody (αMIF). Statistical comparison was performed by one-way ANOVA and corrected using Bonferroni's multiple comparisons test. ns: non-significant.

**Supplementary Table 1. Characteristics of COVID-19 Patients and healthy subjects.**

| | |
|---|---|
| **COVID-19 patients (number)** | 12 |
| Age (Years) | Median=34 (IQR=14) |
| Gender | |
| Male (%) | 90.91% (10/12) |
| Female (%) | 18.18% (2/12) |
| Residency | |
| New Delhi (India) | 100% (12/12) |
| SARS-CoV-2-PCR Positivity | 100% (12/12) |
| Disease Severity* | |
| Mild | 100% (12/12) |
| Symptoms | |
| Fever | 91.67% (11/12) |
| Cough | 75% (9/12) |
| Sore throat | 75% (9/12) |
| Fatigue/Malaise | 58.33% (7/12) |
| Loss of taste | 58.33% (7/12) |
| Loss of smell | 50% (6/12) |
| Diarrhoea | 8.33% (1/12) |
| Days post diagnosis at collection | Median = 90 (IQR=13) |
| **Healthy blood donors (number)** | 26 |
| Age (Years) | Median=31 (IQR=5) |
| Gender | |
| Male (%) | 100% (26/26) |

*WHO Criteria
